# Considerations for implementing a machine learning model for suicide risk in the health systems that serve American Indian communities

**DOI:** 10.64898/2026.10.05.26364799

**Authors:** Rohan R. Dayal, Novalene Alsenay Goklish, Roy Adams, Paul Rebman, Rosemarie Suttle, Mitchell Garcia, Joseph Mega, Valentín Quiroz de la Sierra, Emily E. Haroz

## Abstract

**Objective:** We explored the implementation considerations of machine learning (ML) suicide risk identification in a health system that serves primarily American Indian patients in the Southwestern United States. Previous work developed and reported the performance of suicide risk models, but traditional performance metrics do not directly reflect clinical workflows.

**Materials and Methods:** Using EHR data collected for Emergency Department (ED) visits between 10/01/2019 and 10/02/2021, we conducted Decision Curve Analysis (DCA) to identify the standardized net benefit (SNB) of all models previously evaluated and plotted the potential workload implications of the model with the greatest SNB. Finally, we evaluated suicide screening practices during the study period to inform implementation barriers.

**Results:** Among 9,244 patients, 139 (1.5%) had a suicide attempt or death within 90 days of an included visit. The population-specific logistic regression model had the greatest SNB compared to other approaches. When operationalized, the 95^th^ percentile cutoff did not substantially increase clinical workloads, while the 75^th^ percentile cutoff substantially impacted workloads. The secondary analysis on suicide screening depicted variations in screening administration over time.

**Discussion:** While DCA identified advantages of ML suicide risk identification over current standards, the complexities of suicide-related care limited its clinical interpretability. A stepped-care framework may mitigate potential workload burdens. Training initiatives and dynamic implementation strategies will support high-quality implementation of ML suicide risk identification.

**Conclusion:** Our analyses support the feasibility and utility of augmenting existing suicide screening practices with ML suicide risk identification, and demonstrate the importance of nuanced analyses to inform implementation considerations.

## OBJECTIVE

Almost 49,000 people died by suicide in 2024 in the United States – about 1 death every 11 minutes.[1] Despite slight decreases in recent years, national age-adjusted suicide rates have increased 24.5% over the past 20 years, demonstrating a need for increased prevention efforts.[1] In 2023, non-Hispanic American Indian and Alaska Native (AI/AN) populations had the highest rate of suicide among all racial or ethnic groups in the United States.[2] Differences in suicide rates among AI/AN populations are driven by multilevel factors including historical and intergenerational trauma, inaccessible mental health services, and interpersonal conflict.[3] High endemic rates and recent disproportionate increases in suicide rates among AI/AN communities present a major public health priority.

A key to many public health approaches to suicide prevention is identifying people at risk of suicide. However, this is challenging, particularly in AI/AN communities, as validation studies have not verified that standard risk identification methods are accurate, reliable, and appropriate to the cultures and contexts despite the substantial inequities. For example, despite wide-scale recognition that screening is critical to suicide prevention efforts, there are currently no tools that have been validated specifically with AI/AN populations.[4] Further, existing screening tools primarily identify patients at imminent risk of suicide but are not specifically designed to estimate future risk. Machine learning methods have emerged as a potential solution in improving suicide risk identification by digesting and interpreting vast quantities of data. Several large health systems, including the Veteran’s Administration and private facilities,[5–7] are leveraging these tools to improve care for those at risk of suicide. Recent work has successfully applied ML methods in partnership with AI/AN communities. Shaw and colleagues demonstrated the validity of an existing model for use in a health system that primarily serves AN people.[8] Similarly, our work has demonstrated the utility of these tools outside the health system,[9] the validity of external models in AI populations,[10] the potential added benefit of re-training models specific to certain population demographic characteristics,[11] and strategies of integrating models with suicide screening.[12]

The development and deployment of such models, particularly in resource-constrained settings, comes with a myriad of challenges. First, suicide is a rare event, meaning that any identification tool may have a high false positive rate and result in inappropriate application of limited resources and clinical care. Second, interventions for suicide prevention are limited, with many lacking demonstrated efficacy across a variety of cross-cultural populations, especially among AI/AN populations.[13–14] Finally, in the context of psychiatric care, implementation efforts that necessitate safety measures like involuntary hospitalization must be done carefully and in a way that ensures continued help-seeking and trust in the medical system.[15–17]

Building off prior suicide prevention efforts,[18–19] the Johns Hopkins Center for Indigenous Health, in partnership with several Tribal communities, has developed the Native-Risk Identification for Suicide and Enhanced Care (Native-RISE) system of care. Native-RISE is a systems-level suicide prevention strategy that utilizes ML risk identification combined with brief contact interventions delivered by local case managers to identify patients at risk, facilitate linkages to and engagement with care, provide preventive interventions (e.g., safety planning), and augment existing screening tools used by the Indian Health Service (IHS). The Native-RISE Model (NR-Model) was developed using culturally specific structured Electronic Health Record (EHR) data,[11] and will be used to help identify patients at risk of suicide (e.g., medium or high risk), alert providers, and refer at-risk patients to local case managers. With a goal to implement Native-RISE in three IHS service units in the Southwestern US, the primary provider of health services to over 2.8 million AI/AN patients,[20] careful evaluation of implementation considerations is required.

To inform the development of Native-RISE before its implementation in a Randomized Control Trial, our team aimed to: 1) Examine the clinical utility of ML risk identification through a decision curve analysis (DCA); and 2) Assess the feasibility of Native-RISE by generating workload estimates of current suicide screening practices augmented by the NR-Model as an additional risk identification method, and examining the implementation of universal suicide screening at Emergency Department (ED) visits in this care setting. The results of this study will provide critical local insights and illustrate important considerations for studies to examine when implementing ML models in clinical care. Further, recommendations from these results will provide novel findings to the growing field of research utilizing ML methods to reduce health disparities.

## MATERIALS AND METHODS

This manuscript adheres to the reporting guidelines established in the Strengthening the Reporting of Observational studies in Epidemiology (STROBE) Statement.[21]

### Ethical and Tribal Approvals

This study builds on a 40+ year relationship between Tribal partners and the Johns Hopkins Center for Indigenous Health. This study was conducted in accordance with the Declaration of Helsinki, and was approved by the Phoenix Area Office Institutional Review Board (IRB) of the Indian Health Service and the Johns Hopkins Bloomberg School of Public Health IRBs (PXR 22.05; JHSPH 17333). A waiver of consent was granted by the IHS IRB through a HIPAA authorization to conduct this secondary analysis of existing EHR data. The White Mountain Apache Tribe (WMAT) approved the study through a Tribal Resolution. All analyses of protected health information were conducted in a secure computing environment administered by the Johns Hopkins School of Medicine.

### Population

We previously conducted cohort analyses of EHR data for patients age 18 and older who had clinical encounters at the IHS-operated Whiteriver Service Unit (WRSU) between January 1, 2017, and October 2, 2021, to test existing and develop new ML models for suicide risk identification.[10–11] The WRSU provides inpatient, outpatient, pharmacy, and emergency care to approximately 17,000 Tribal members, in addition to non-Tribal members in the surrounding communities. In these analyses, suicide risk probabilities were generated for 331,588 visits between 1/1/2017 and 10/2/2021 that were considered actionable for patient intervention (i.e. primary care, emergency department, mental health-related visits, etc.).

In the fall of 2019, a mandate was established at the WRSU to screen all patients presenting to the Emergency Department (ED) using the Ask Suicide Questions (ASQ) Survey.[22]. This current study employed the previous cohort and filtered to Emergency Department (ED) visits between October 1, 2019, and October 2, 2021 to better draw comparisons between ML risk identification and current standard practices. As a sensitivity analysis, we repeated the analyses on all visits with a probability of suicide risk within the same date range to inform implementation considerations across all departments of the service unit (Supplemental File 1).

### Models

Full details on model development, feature inclusion, training, and testing are described in greater detail elsewhere.[10–11] Briefly, we incorporated two pre-existing in this study: a random forest model developed at the Vanderbilt University Medical Center (“VU model”),[23] and an outpatient mental health logistic regression model developed by the Mental Health Research Network (“MHRN model”).[24] Previously with this cohort, we reported that the MHRN model had higher discriminative ability than the VU model at identifying past 90-day attempts (Area under the Receive Operating Curve [AUROC] for the MHRN model = 0.81; AUROC for the VU model = 0.68).[10]

Two models developed by our team in previous analyses were incorporated as well: a random forest classification model (“IHS-RF model’’), and a logistic model (“IHS-LR model”). Both were re-trained versions of the MHRN model with locally relevant features (i.e. diagnosis codes for substance use, intimate partner violence screens, etc.).[11] In our previous reporting, both models had equally high discriminative ability (AUROC = 0.83) in identifying past 90-day attempts or deaths.

Finally, we operationalized an enhanced screening metric (“existing screening”), as used in our previous reporting, which included: 1) positive screening on the ASQ or the Columbia-Suicide Severity Rating Scale (C-SSRS),[22, 25] 2) diagnosis of suicidal ideation in the past 90 days, or 3) any prior suicide attempt.[10–11] The existing screening metric included all possible tools clinicians use to identify suicide risk, and had an AUROC of 0.66 in identifying past 90-day suicide attempts.[10]

For the purposes of this current study, we utilized the outcome of 90-day suicide attempts or deaths for all analyses. All predicted probabilities were recalibrated to this outcome using isotonic regression to avoid miscalibration (Supplemental File 2).

### Net benefit

We conducted a Decision Curve Analysis (DCA) to quantify the clinical usefulness of all models and existing screening. Decision curves plot net benefit, a metric that estimates the clinical benefit of a model through the subtraction of the relative value of false-positive and true– positive classifications, against discrete probability thresholds.[26–27] DCA is advantageous in cases when data on harms and benefits are difficult. Net benefit is plotted across a range of discrete thresholds to approximate the threshold probability of a model, or the point at which the expected benefits and potential harms of treatment are equal.[26–27] Decision curves also consider both model discrimination and calibration, giving a more holistic view of model performance.[28] Therefore, while some models may have a higher overall accuracy, a model may have a lower net benefit at the desired cutoff.

Decision curves were plotted for all four models described above, existing screening, and two additional treatment scenarios: 1) all patients are considered at risk and are treated (“Treat All”), and 2) no patients are considered at risk or are treated for a potential suicide outcome (“Treat None”). The treat all and treat none approaches were included to determine whether the models had any clinical utility at a given threshold. Standardized Net Benefit (SNB), a metric estimating net benefit standardized by outcome prevalence, with a range of 0-100%, was used to make comparisons across models.[28] To improve the accuracy of the decision curves and all reported summary statistics, estimates were averaged across the 10 cross-validated folds created at the patient-level during model development.[11] R V4.3.1 and RStudio were used to quantify SNB with the “dcurves” package.[29–30] Additional details and example interpretations related to net benefit are included in Supplemental File 3.

### Estimating Impact on Workloads

To better inform feasibility, we examined the potential added workload arising from risk alerts generated by the NR-Model. We leveraged the IHS-LR model-recalibrated predicted probabilities, and operationalized discrete cutoffs. These cutoffs were informed through discussions with providers, who stated substantial increases to the number of patients identified with suicide risk would be challenging given limited existing personnel, highly geographically isolated care settings, and alert fatigue. Providers also indicated a desire for multiple categories of risk to implement risk-concordant care.

The medium and high-risk cutoffs were set at the 95th and 75^th^ percentile of all predicted probabilities, respectively, to achieve sensitivity values of approximately 40% and 80%. The medium and high-risk groups were designated to be mutually exclusive. Given that risk scores aim to identify risk over 90 days and that people at risk might visit the ED more than once in that 90-day window, we limited alerts to cases where patients transitioned from medium to high, low to medium, and stayed at medium or high risk with no visit in the past 90 days. Finally, we generated weekly workload estimates for patients who were identified as at-risk via the NR-Model or existing screening to understand how the implementation of risk alerts would impact current workloads.

### Universal ASQ Screening Implementation

Given that the dataset spanned the initial implementation of universal suicide risk screening in the ED, we examined the percentage of visits with a recorded ASQ screening result over time to understand potential implementation barriers for Native-RISE. To assess the administration of ASQs over time, we calculated the percentage of ED visits with a recorded positive or non-acute positive ASQ result per month. RStudio V4.3.1 packages “dplyr” and “plotly”,[31–32] respectively, were used to estimate and visualize all figures.

## RESULTS

### Descriptive Statistics

The overall sample of ED visits included 9,244 patients with 42,915 ED visits between 10/1/2019 and 10/2/2021. Of these patients, the median age at first visit was 38 (IQR: 28, 54), and 52.7% of patients were Female. Almost all patients identified their ethnicity as not Hispanic, Latino, or Spanish origin (98.5%) and their race as American Indian or Alaska Native (97.1%). One hundred thirty-nine (1.5%) patients had a documented suicide attempt or death due to suicide, and 78 (0.8%) patients had a documented attempt or death within 90 days of an included ED visit. Approximately one-tenth of all patients screened positive for suicidality at one or more of their visits through existing screening (N = 945, 10.2%) (**Table 1**).

**Table 1.**
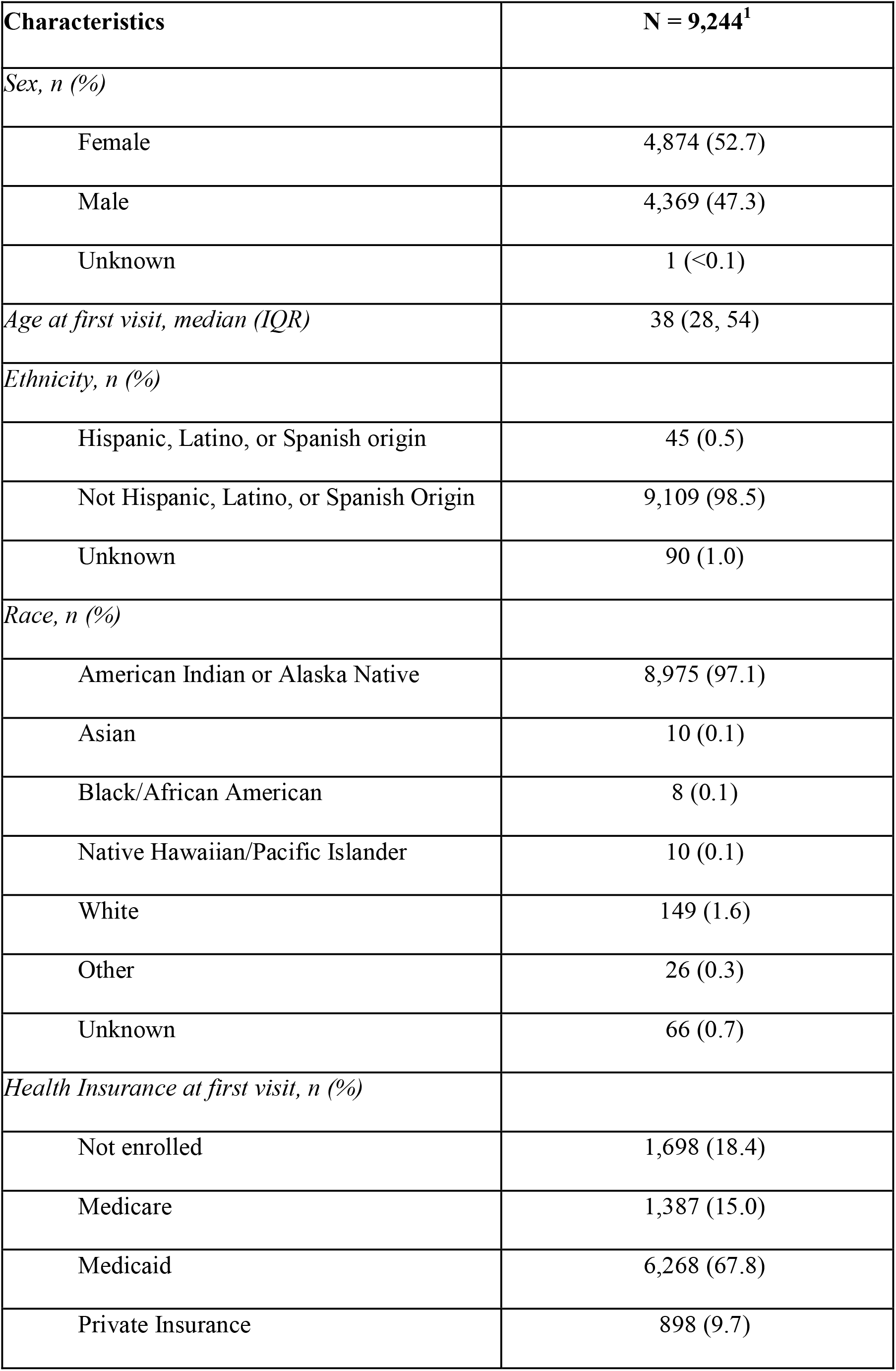
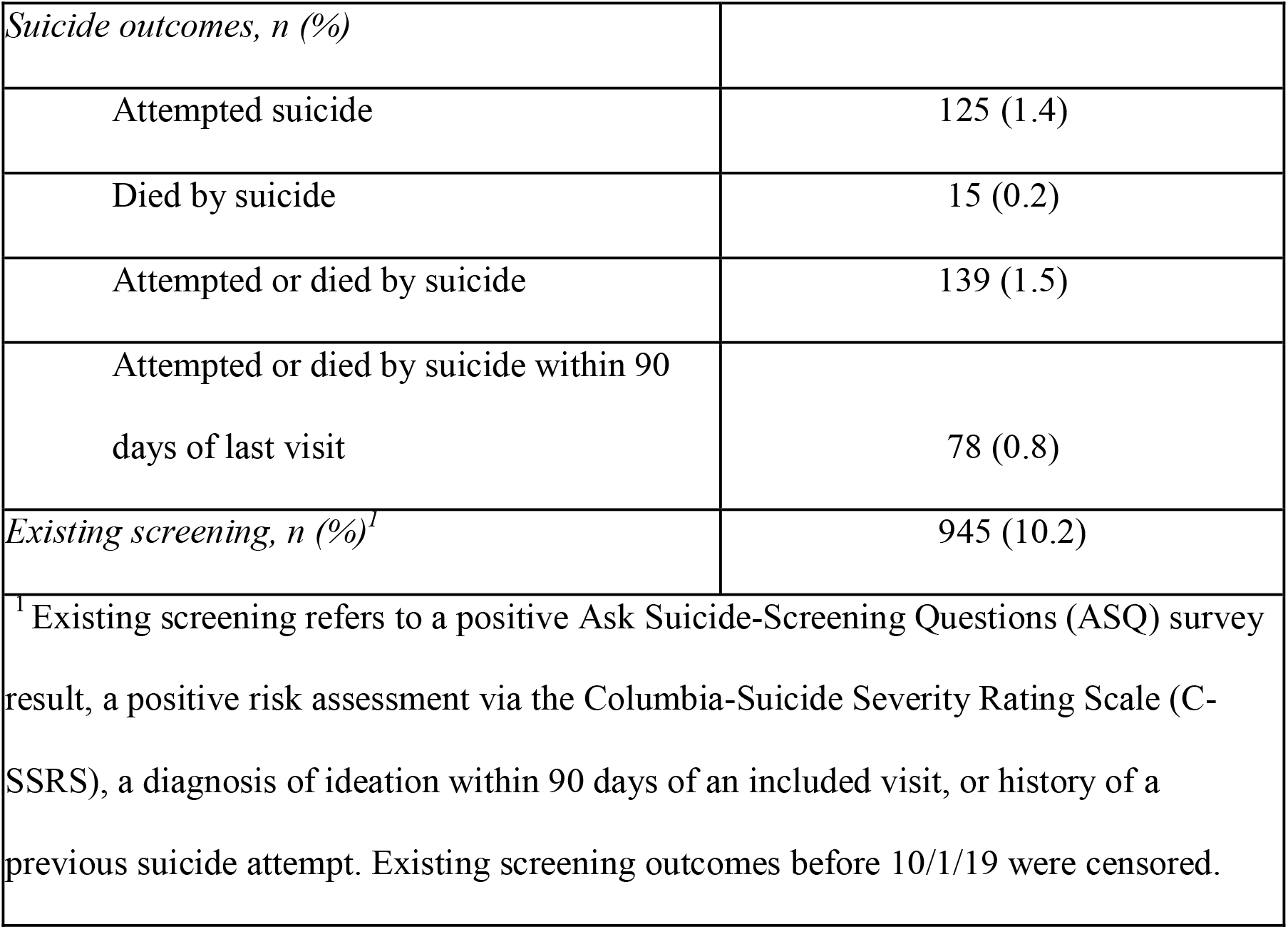
– Sample Demographic Characteristics of Patients with One or More Emergency Department (ED) Visits Between October 2019 and October 2021 (N = 9,244 patients; 42,915 visits)

| Characteristics | N = 9,244<sup>1</sup> |
| --- | --- |
| *Sex, n (%)* |  |
| Female | 4,874 (52.7) |
| Male | 4,369 (47.3) |
| Unknown | 1 (<0.1) |
| *Age at first visit, median (IQR)* | 38 (28, 54) |
| *Ethnicity, n (%)* |  |
| Hispanic, Latino, or Spanish origin | 45 (0.5) |
| Not Hispanic, Latino, or Spanish Origin | 9,109 (98.5) |
| Unknown | 90 (1.0) |
| *Race, n (%)* |  |
| American Indian or Alaska Native | 8,975 (97.1) |
| Asian | 10 (0.1) |
| Black/African American | 8 (0.1) |
| Native Hawaiian/Pacific Islander | 10 (0.1) |
| White | 149 (1.6) |
| Other | 26 (0.3) |
| Unknown | 66 (0.7) |
| *Health Insurance at first visit, n (%)* |  |
| Not enrolled | 1,698 (18.4) |
| Medicare | 1,387 (15.0) |
| Medicaid | 6,268 (67.8) |
| Private Insurance | 898 (9.7) |
| *Suicide outcomes, n (%)* |  |
| Attempted suicide | 125 (1.4) |
| Died by suicide | 15 (0.2) |
| Attempted or died by suicide | 139 (1.5) |
| Attempted or died by suicide within 90 days of last visit | 78 (0.8) |
| *Existing screening, n (%)*<sup>1</sup> | 945 (10.2) |
| <sup>1</sup> Existing screening refers to a positive Ask Suicide-Screening Questions (ASQ) survey result, a positive risk assessment via the Columbia-Suicide Severity Rating Scale (C-SSRS), a diagnosis of ideation within 90 days of an included visit, or history of a previous suicide attempt. Existing screening outcomes before 10/1/19 were censored. |  |

### Net Benefit

Figure 1 compares the net benefits of existing screening, and the VU, MHRN, IHS-LR model, and IHS-RF models in identifying 90-day suicide attempts or deaths. Existing screening and the MHRN, IHS-RF, and IHS-LR models were justified for clinical use at probability thresholds below approximately 3%. The VU model reflected the lowest SNB of all included models in this sample, and was not justifiable for clinical use due to having equivalent SNB values to the Treat All curve below probability thresholds of approximately 0.3% and SNB values equivalent to the Treat None curve above probability thresholds of approximately 1.4%.

**Figure 1.**
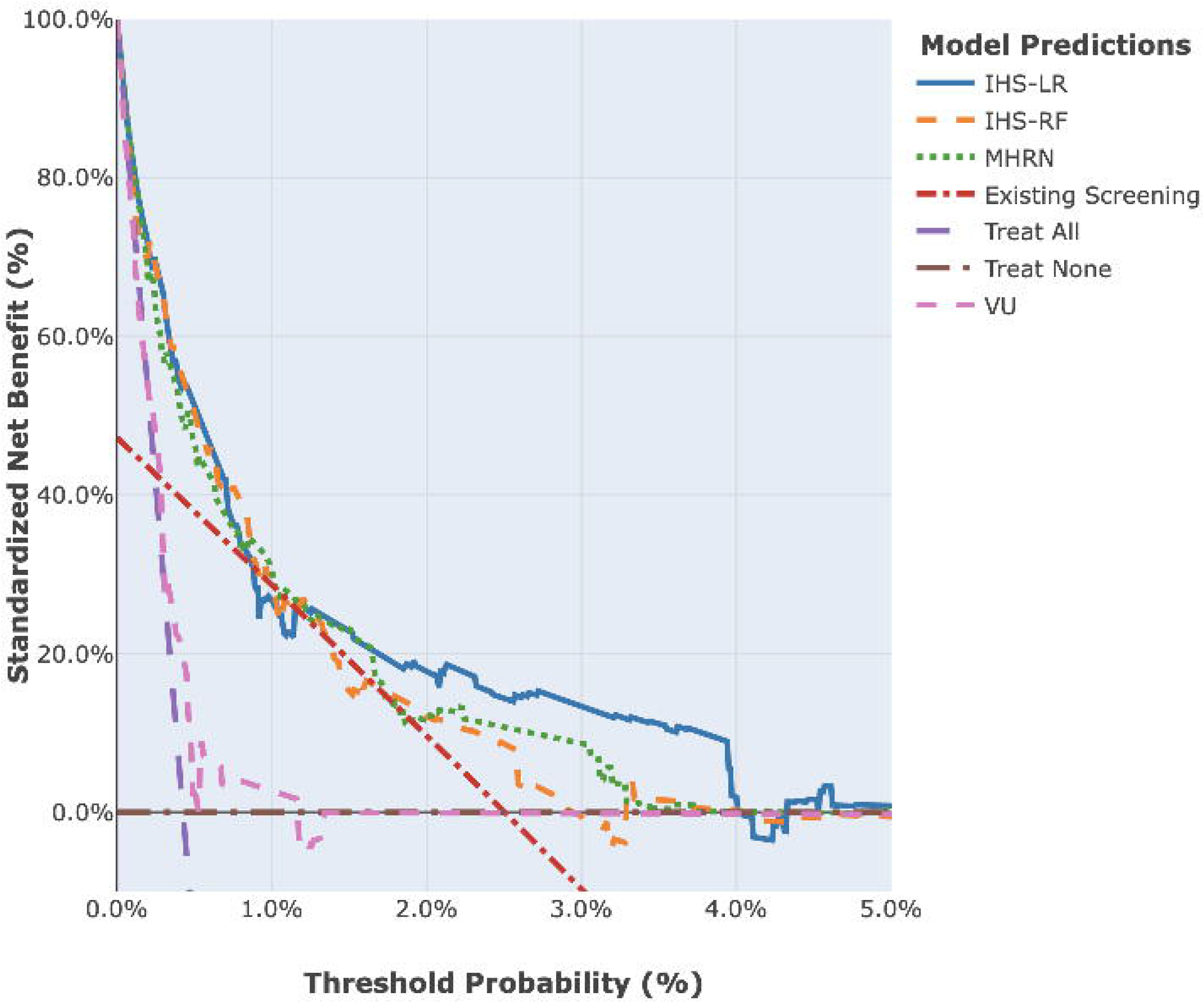
– Comparison of Standardized Net Benefits for the Identification of Patients Who Had a Suicide Attempt or Died by Suicide Within 90 Days of Emergency Department (ED) Visits Between October 2019 and October 2021 (N = 9,244 patients; 42,915 visits) *Alt Text:* Decision curves for the Vanderbilt University (VU), Mental Health Research Network (MHRN), IHS logistic regression (LR), and IHS random forest (RF) models, as well as existing screening, plotting standardized net benefit (0-100%) against threshold probabilities between 0% and 5%.

In comparison to existing screening, the MHRN, IHS-RF, and IHS-LR models all had greater SNB values at thresholds below approximately 0.9%. While the MHRN and IHS-RF models had SNB values equivalent to or slightly greater than existing screening and the Treat None curve at thresholds between 1.2% and 3.5%, the IHS-LR model’s decision curve was greater than all approaches at thresholds between 1.2% and 4%.

### Operationalizing the IHS-LR Model as the Native-RISE Model

The high-risk cutoff, operationalized at the 95th percentile of predicted probabilities, had a sensitivity of 42.6%, specificity of 93.7%, positive predictive value (PPV) of 3.19%, false positive rate (FPR) of 6.24%, and false negative rate (FNR) of 0.26%. The medium-risk cutoff, operationalized at the 75th percentile (thus including both medium– and high-risk classifications), had a sensitivity of 85.5%, specificity of 69.3%, PPV of 1.28%, FPR of 30.6%, and FNR of 0.07%. However, when limited to the medium-risk category, sensitivity dropped to 38.9% and specificity to 79.0%. In comparison, existing screening had a sensitivity of 47.2%, specificity of 91.4%, PPV of 2.50%, FPR of 8.52%, and FNR of 0.24%. During the study period, 426 (4.6%) patients were flagged as high risk for at least one visit, and 1,893 (20.5%) were flagged as medium risk for at least one visit. **Table 2** reflects the comparison of SNB between the IHS-LR model and existing screening at cutoffs ranging from the 50th to the 99th percentiles of IHS-LR model predicted probabilities.

**Table 2.**
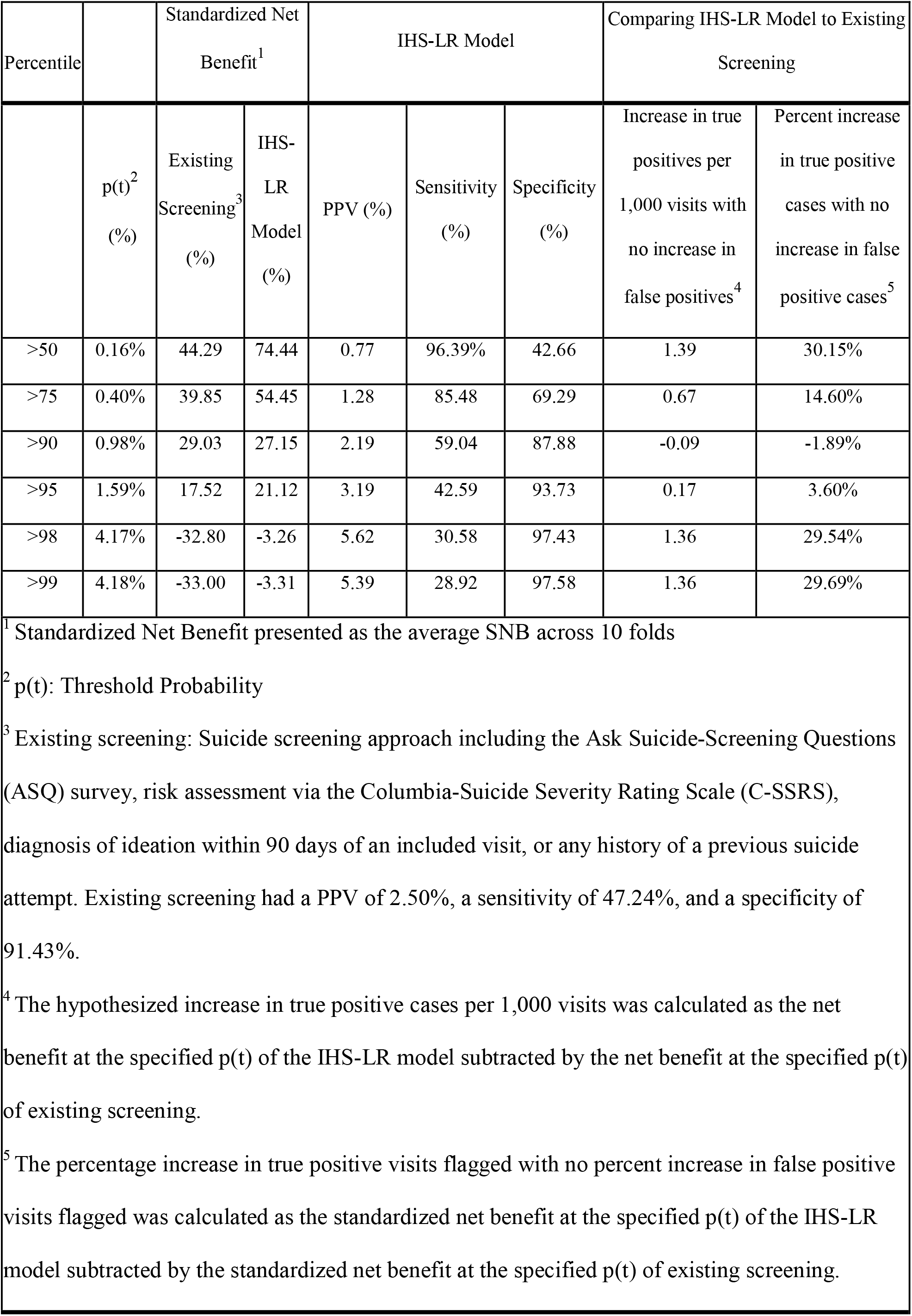
– Standardized Net Benefit, Summary Statistics, and Predicted Advantages of the IHS Logistic Regression (IHS-LR) Model in Comparison to Existing Screening Among Emergency Department (ED) Visits Between October 2019 and October 2021 (N = 9,244 patients; 42,915 visits)

Comparing the NR-Model medium-risk category to existing screening resulted in an increase of approximately 0.7 more correctly identified visits with at-risk patients per 1,000 visits, or a 14.6% increase, with no increase in the number of incorrectly identified visits. The high-risk category identified approximately 0.2 more correctly identified visits with at-risk patients per 1,000 visits, or a 3.6% increase, with no increase in the number of incorrectly identified visits compared to existing screening.

### Estimating Impact on Workloads

During the study period, the median number of visits per week in the ED was 414 (IQR: 350, 482). Based on the augmented existing screening approach, clinicians encountered a patient who screened positive for suicide at a median 8.9% (IQR: 7.0%, 10.4%) of ED visits per week (**Figure 2**). The NR-Model high-risk classification would have identified a median 1.3% (IQR: 0.9%, 1.7%) of visits per week, while the median number of visits flagged by the medium-risk category per week was 6.8% (IQR: 5.9%, 7.7%). Combining the NR-Model with existing screening, the high-risk flag increased the median percentage of visits per week identified from 8.9% to 10.0% (IQR: 8.2%, 12.0%), while the combined medium and high-risk flag increased the median percentage of weekly identified visits to 15.0% (IQR: 13.0%, 17.6%).

**Figure 2.**
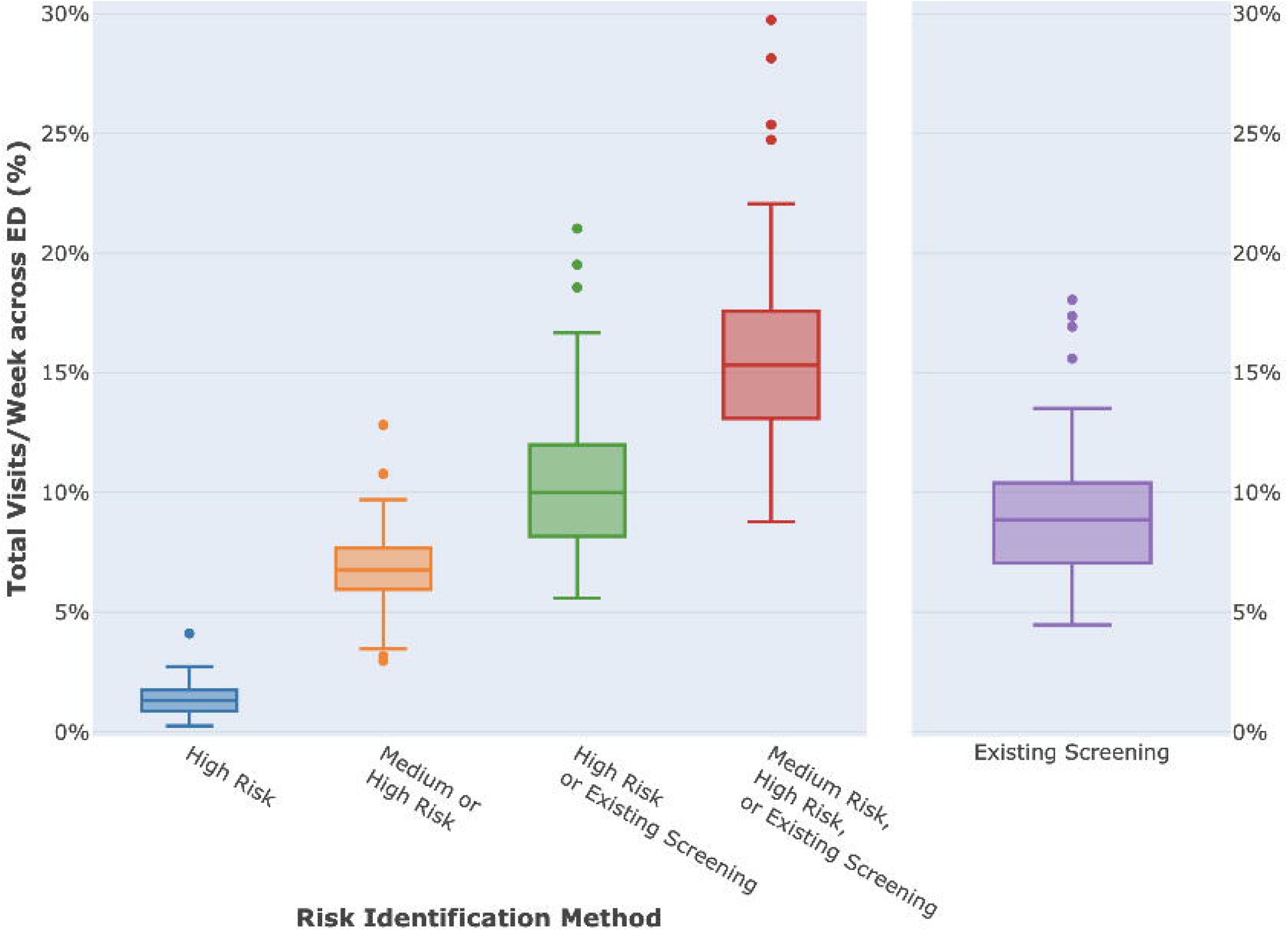
– Workload Estimates in Percentage of Alerts on Emergency Department (ED) Visits per Week Comparing the Native-RISE IHS-LR Model to Existing Screening Between October 2019 and October 2021 (N = 9,244 patients; 42,915 visits) *Alt Text:* Graphical depiction of weekly workload estimates for the Emergency Department of this IHS Service Unit upon implementing the IHS logistic regression (LR) model and operationalizing it at the 95^th^ and 75^th^ percentiles for high and medium risk categories, respectively.

### Universal ASQ Screening Implementation

We found changes over time in ASQ screening practices for visits that occurred in the ED (**Figure 3**). Shortly after universal suicide screening using the ASQ was implemented, there was a sharp increase in the percentage of visits that were screened to a median of approximately 90% in February 2020. The period between February 2020 and September 2021 saw gradual decreases from a peak of approximately 90% to a low of approximately 53%. The overall percentage of visits screened for suicidality for ED-only visits across the entire study duration was 62.7%.

**Figure 3.**
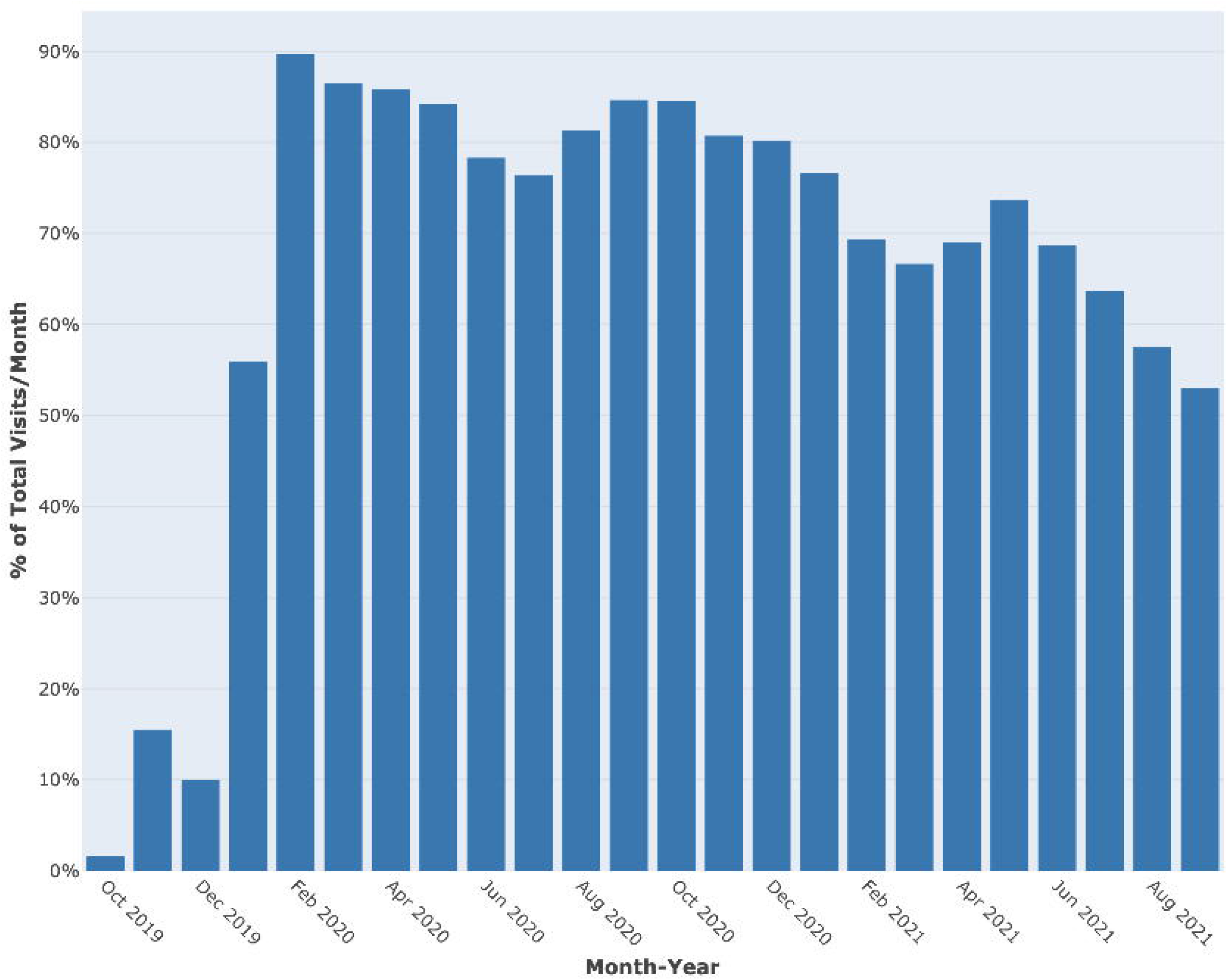
– Percentage of Monthly Emergency Department (ED) Visits with an Administered Ask Suicide-Screening Questions (ASQ) Survey Between October 2019 and October 2021 (N = 9,244 patients; 42,915 visits) *Alt Text:* Histogram of monthly screening rates with the ASQ survey upon implementation of a universal suicide screening mandate at this IHS service unit.

## DISCUSSION

The logistic regression model retrained with locally relevant data (IHS-LR model) presented the greatest overall clinical utility. Our hypothesized workload estimates identified minimal additional workload burden for high risk identifications and greater workload increases for medium risk identifications, highlighting the importance of risk-concordant care. Lastly, the variation observed in ASQ screening practices over time has important implications on potential implementation barriers. These findings support the feasibility of using the NR-Model as an addition to existing screening practices to enhance suicide risk identification. The sensitivity analyses further highlight that the utility and feasibility of the NR-Model may extend to non-acute care settings as well.

### Clinical Utility

While criticisms of suicide risk ML models have argued that models may not yield actionable results due to having low PPVs,[33] alternative methods like DCA, cost-effectiveness analyses, and workload assessments have been proposed to demonstrate that low PPVs may be tolerated to improve risk identification for rare and severe outcomes.[34–36] Plotting the SNB of all models revealed that the IHS-LR model outperformed all approaches after equalizing for false positive identifications and recalibrating probabilities, and identified more actionable true positive cases compared to the IHS-RF model despite their equivalent discriminative ability (AUROC = 0.83).[11] However, the advantage of the IHS-LR model over existing screening was marginal at thresholds between 0.9% and 1.2%, suggesting that the model’s strength lies primarily in identifying those at exceedingly high risk or ruling out those at low risk.

Despite the advantages of DCA, the complexities of suicide risk identification and suicide-related care in this setting limited the clinical interpretability of DCA. The consequences of identifying and treating suicide risk in this context vary considerably by individual circumstances – including suicide risk severity as determined by a formal suicide risk assessment, workload capacity for admission, and consultation resources. Moreover, the benefits extend beyond immediate clinical care. In our setting, identification of suicide risk also triggers community-based case management referrals, which have demonstrated high success rates at reducing suicide outcomes.[19] Variability in the operationalization of treatment for suicide risk corresponded with variability in the interpretation of net benefit.

Future studies should explore the net benefits of such models in more detail, including having clinicians specifically consider different types of potential treatments or interventions similar to the modeling approach taken by McCutchan et al.[37] Future studies should also consider exploring the impact of financial costs incurred through the implementation of suicide risk models on net benefit.

### Workload Feasibility

To address provider concerns for increasing workloads and gain clinician buy-in for the NR-Model, we analyzed the alert burden of the NR-Model compared to existing practices. Our findings suggest minimal additional alert burden for the integration of the NR-Model high risk classification with existing screening (median visits per week: 10.0%) compared to existing screening alone (median visits per week: 8.9%). The medium-risk combined with existing screening increased alert frequency more substantially (median visits per week: 15.0%), but this category serves an important purpose in boosting identification sensitivity and facilitating risk-concordant care in a highly acute population where many patients fall into this intermediate range.

While some argue that discretizing probabilities from risk models limits the ability of clinicians to make personalized treatment decisions,[35,38] a risk-concordant stepped-care approach is essential in ensuring appropriate resource allocation and promoting actionable workloads. The NR-Model will be implemented in an environment where high-risk suicide care requires immense allocation of limited resources (e.g., emergency transport over 300 miles to an inpatient treatment facility). Health systems implementing suicide risk identification models, especially those with limited resources, should carefully consider operational capacity and treatment options within a stepped-care framework to mitigate potential implementation barriers.

In a pre-implementation feasibility study, Kline-Simon et al. found that alerting on every visit at or above the 95^th^ percentile for suicide risk resulted in a median of 162 alerts per day (sensitivity = 41.3%, PPV = 6.4%), while alerting on patients’ first at-risk visit reduced alerts to a median of 2 per day.[36] Similarly, we sought to reduce alert fatigue by excluding visits that were identified through existing screening, and limiting alerts to visits where a patient’s risk category increased (e.g., medium to high risk) or remained high despite not seeking care for an extended period. Because suicide screening identifies a substantial number of patients and requires extensive resource allocation, integrating NR-Model alerts into existing screening practices provides an avenue to mitigate alert fatigue that preserves existing workflows. Ultimately, model optimization relies not just on its performance, but on the adopting health system’s capacity.

### Implementation Considerations

Our evaluation of the implementation of universal suicide screening at ED visits found an initial peak in ASQ survey administration, followed by a gradual decline throughout the remainder of the study duration. While the exact reason for this decline is unclear, this trend must be discussed within the overall context of this setting. Variations in ASQ screening over time may result from high staff turnover and a large reliance on locum providers, both of which impact this rural IHS facility and many others. Additionally, this implementation effort may have been impeded by the COVID-19 pandemic, which introduced additional constraints and reduced patient access to non-COVID-related care. Studies have argued that barriers to universal suicide screening efforts can be overcome by proper training initiatives or staff reallocations but also acknowledge that adoption inherently increases workload burdens.[39] Existing literature and observed ASQ screening trends indicate that quality improvement approaches, such as training and training booster sessions, will be necessary to support the high-quality implementation of Native-RISE. Additionally, implementation strategies must be dynamic to account for time-specific health system constraints or ML-specific considerations, such as model drift, to ensure success over time.

### Limitations

This study should be interpreted within the context of its limitations. First, suicide-related outcomes that were not reported (i.e. miscoded in the EHR, did not require medical attention or were not disclosed, treated at other facilities, etc.) in the data may result in underestimates of SNB and other model summary statistics. Second, the interpretability of the DCA may be limited due to the difficulty in identifying a feasible range of probability thresholds given the considerable variation in treatment consequences. Third, we grouped acute and non-acute positive results on the ASQ to estimate the number of visits with any suicide risk identified. However, as providers are trained to respond to acute and non-acute ASQ positive responses differently, this inclusion potentially overestimates the number of visits that would require additional suicide risk assessment and further complicates the understanding of existing screening workloads. Lastly, the workload estimates may not generalize the workload implications of suicide risk models in other healthcare systems.

## CONCLUSION

Our analysis is the first to examine clinical utility and workload implications of a suicide risk identification model in a rural American Indian population. Previous work has demonstrated the potential for ML models to reify or uphold existing problematic patterns in data.[40] When developed in partnership with Tribal communities and incorporating local knowledge, ML models can potentially improve access to healthcare services and overall quality of care, and reduce adverse health outcomes for AI/AN people. This approach, incorporated throughout the development, training, validation, and implementation of Native-RISE, challenges the notion of one-size-fits-all solutions by instead offering a framework to integrating models that are deeply rooted in the contexts of specific communities.

The Native-RISE Model, chosen as the IHS-specific logistic regression model, had a greater SNB than all other models tested throughout all plausible threshold probability ranges. Our workload estimates indicated that while high-risk visits identified by the model would not result in a substantial increase in workload, additional considerations are needed for the successful implementation of a medium-risk category. Model performance and evaluation must extend traditional statistical measures to include clinical and implementation considerations. In the context of current practices and the health system, nuanced reporting of implementation considerations is critical to advancing our understanding of how these models can better serve communities.

## Declarations

### Ethics approval and consent to participate

This study was conducted in accordance with the Declaration of Helsinki, and all study procedures were approved by the Phoenix Area Office Institutional Review Board of the Indian Health Service and the Johns Hopkins Bloomberg School of Public Health Institutional Review Boards (PXR 22.05; JHSPH 17333). Local approvals from the White Mountain Apache Tribe (WMAT) were provided through a Tribal Resolution. The data for this secondary analysis of EHR data was accessed through a data use agreement between the Indian Health Service and Johns Hopkins University. Informed consent was waived by a HIPAA authorization granted by the PXR Institutional Review Board in accordance with 45 CFR 46.116(f).

### Consent for publication

Not applicable.

### Clinical trial number

Not applicable.

### Availability of data and materials

The data that support the findings of this study are available from the White Mountain Apache Tribe and Indian Health Service, but restrictions apply to the availability of these data, which were accessed under a data use agreement for the current study, and so are not publicly available. Data are however available from the authors upon reasonable request and with permission of the White Mountain Apache Tribe and Indian Health Service.

### Authors’ contributions

Rohan R. Dayal: conceptualization, data curation, formal analysis, methodology, project administration, visualization, writing – original draft preparation and reviewing & editing; Novalene Alsenay Goklish: conceptualization, funding acquisition, investigation, writing – original draft preparation and reviewing & editing; Roy Adams: data curation, investigation, methodology, resources, software, supervision, writing – original draft preparation and reviewing & editing; Paul Rebman: data curation, methodology, writing – reviewing & editing; Rosemarie Suttle: writing – reviewing & editing; Mitchell Garcia: writing – reviewing & editing; Jospeh Mega: writing – reviewing & editing; Valentín Quiroz de la Sierra: writing – original draft preparation; Emily E. Haroz: conceptualization, funding acquisition, investigation, methodology, resources, supervision, validation, writing – original draft preparation and reviewing & editing.

## List of Abbreviations

AI/AN: American Indian and Alaska Native
ML: Machine Learning
Native-RISE: Native-Risk Identification for Suicide and Enhanced Care
NR-Model: Native-RISE Model
IHS: Indian Health Service
DCA: Decision Curve Analysis
WMAT: White Mountain Apache Tribe
EHR: Electronic Health Record
WRSU: White River Service Unit
ED: Emergency Department
VU: Vanderbilt University
MHRN: Mental Health Research Network
ASQ: Ask Suicide Questions
AUROC: Area Under the Receiver Operating Curve
IHS-RF Model: Indian Health Service Random Forest Model
IHS-LR Model: Indian Health Service Logistic Regression Model
SNB: Standardized Net Benefit
PPV: Positive Predictive Value
IQR: Interquartile Range

## Acknowledgements

We humbly acknowledge the White Mountain Apache Tribe and its community members for their support in undertaking this project. We continue to strive to help promote life for all in the community and elsewhere. We also thank Dr. Gregory Simon and the team at Kaiser Permanente, as well as Dr. Colin Walsh and his team, for providing their open-source code, ongoing guidance, and feedback. While the study was done in collaboration with the Indian Health Service, the contents do not represent the views of the Indian Health Service or the United States Government.

## REFERENCES

1. Web-Based Injury Statistics Query and Reporting System (WISQARS). National Center for Injury Prevention and Control, Centers for Disease Control and Prevention (CDC). https://www.cdc.gov/injury/wisqars/index.html (accessed 24 Sept, 2026).

2. Stone DM, Cammack AL, Carbone EG. Notes from the Field: Differences in Suicide Rates, by Race and Ethnicity and Age Group — United States, 2018-2023. MMWR Morb Mortal Wkly Rep 2025;74:550–553. doi: 10.15585/mmwr.mm7435a2.

3. Stone D, Trinh E, Zhou H, et al. Suicides Among American Indian or Alaska Native Persons — National Violent Death Reporting System, United States, 2015–2020. MMWR Morb Mortal Wkly Rep. 2022;71:1161–8. doi: 10.15585/mmwr.mm7137a1

4. Shaw, CE. The critical need for a culturally sensitive suicide risk assessment instrument for American Indian and Alaska Native youth [Master’s alternative plan paper, Minnesota State University, Mankato]. Cornerstone: A Collection of Scholarly and Creative Works for Minnesota State University, Mankato. 2021. https://cornerstone.lib.mnsu.edu/etds/1103/ (accessed 24 Sept, 2026).

5. Matarazzo BB, Eagan A, Landes SJ, et al. The Veterans Health Administration REACH VET Program: Suicide Predictive Modeling in Practice. PS. 2023;74:206–9. doi: 10.1176/appi.ps.202100629

6. Richards JE, Yarborough BJH, Holden E, et al. Implementation of Suicide Risk Estimation Analytics to Support Mental Health Care for Quality Improvement. JAMA Netw Open. 2022;5:e2247195. doi: 10.1001/jamanetworkopen.2022.47195

7. Walsh CG, Ripperger MA, Novak L, et al. Risk Model–Guided Clinical Decision Support for Suicide Screening: A Randomized Clinical Trial. JAMA Netw Open. 2025;8:e2452371. doi: 10.1001/jamanetworkopen.2024.52371

8. Shaw JL, Beans JA, Noonan C, et al. Validating a predictive algorithm for suicide risk with Alaska Native populations. Suicide & Life Threat Behav. 2022;52:696–704. doi: 10.1111/sltb.12853

9. Haroz EE, Goklish N, Walsh CG, et al. Evaluation of the Risk Identification for Suicide and Enhanced Care Model in a Native American Community. JAMA Psychiatry. 2023;80:675. doi: 10.1001/jamapsychiatry.2022.5068

10. Haroz EE, Rebman P, Goklish N, et al. Performance of Machine Learning Suicide Risk Models in an American Indian Population. JAMA Netw Open. 2024;7:e2439269. doi: 10.1001/jamanetworkopen.2024.39269

11. Adams R, Haroz EE, Rebman P, et al. Developing a suicide risk model for use in the Indian Health Service. npj Mental Health Res. 2024;3:47. doi: 10.1038/s44184-024-00088-5

12. Goklish NA, Haroz EE, Dayal RR, et al. Combining Machine Learning Models and Screening to Enhance Suicide Risk Identification for American Indian Patients: Retrospective Cohort Study. J Med Internet Res. 2026;28:e82669–e82669. doi: 10.2196/82669

13. Clifford AC, Doran CM, Tsey K. A systematic review of suicide prevention interventions targeting indigenous peoples in Australia, United States, Canada and New Zealand. BMC Public Health. 2013;13:463. doi: 10.1186/1471-2458-13-463

14. Pham TV, Fetter AK, Wiglesworth A, et al. Suicide interventions for American Indian and Alaska Native populations: A systematic review of outcomes. SSM – Mental Health. 2021;1:100029. doi: 10.1016/j.ssmmh.2021.100029

15. Ansloos J. Rethinking Indigenous Suicide. IJIH. 2018;13:8–28. doi: 10.32799/ijih.v13i2.32061

16. Wexler LM, Gone JP. Exploring Possibilities for Indigenous Suicide Prevention. In: White J, Marsh I, Kral MJ, Morris J, editors. Critical suicidology: transforming suicide research and prevention for the 21st century. Vancouver: UBC Press; 2016. p. 56–70.

17. Chrisjohn RD, McKay SM, Smith AO. Dying to please you: Indigenous suicide in contemporary Canada. Penticton, BC: Theytus Books Ltd.; 2017.

18. Cwik MF, Barlow A, Goklish N, et al. Community-Based Surveillance and Case Management for Suicide Prevention: An American Indian Tribally Initiated System. Am J Public Health. 2014;104:e18–23. doi: 10.2105/AJPH.2014.301872

19. Cwik MF, Tingey L, Maschino A, et al. Decreases in Suicide Deaths and Attempts Linked to the White Mountain Apache Suicide Surveillance and Prevention System, 2001–2012. Am J Public Health. 2016;106:2183–9. doi: 10.2105/AJPH.2016.303453

20. IHS Profile. In: Indian Health Service Fact Sheet. 2024. https://www.ihs.gov/sites/newsroom/themes/responsive2017/display_objects/documents/factsheets/IHSProfile.pdf (accessed 24 Sept 2026).

21. Elm EV, Altman DG, Egger M, et al. Strengthening the reporting of observational studies in epidemiology (STROBE) statement: guidelines for reporting observational studies. BMJ. 2007;335:806–8. doi: 10.1136/bmj.39335.541782.AD

22. Horowitz LM, Bridge JA, Teach SJ, et al. Ask Suicide-Screening Questions (ASQ): A Brief Instrument for the Pediatric Emergency Department. Arch Pediatr Adolesc Med. 2012;166:1170. doi: 10.1001/archpediatrics.2012.1276

23. Walsh CG, Johnson KB, Ripperger M, et al. Prospective Validation of an Electronic Health Record–Based, Real-Time Suicide Risk Model. JAMA Netw Open. 2021;4:e211428. doi: 10.1001/jamanetworkopen.2021.1428

24. Simon GE, Johnson E, Lawrence JM, et al. Predicting Suicide Attempts and Suicide Deaths Following Outpatient Visits Using Electronic Health Records. AJP. 2018;175:951–60. doi: 10.1176/appi.ajp.2018.17101167

25. Posner K, Brown GK, Stanley B, et al. The Columbia–Suicide Severity Rating Scale: Initial Validity and Internal Consistency Findings From Three Multisite Studies With Adolescents and Adults. AJP. 2011;168:1266–77. doi: 10.1176/appi.ajp.2011.10111704

26. Steyerberg EW, Vickers AJ, Cook NR, et al. Assessing the Performance of Prediction Models: A Framework for Traditional and Novel Measures. Epidemiology. 2010;21:128–38. doi: 10.1097/EDE.0b013e3181c30fb2

27. Vickers AJ, Elkin EB. Decision Curve Analysis: A Novel Method for Evaluating Prediction Models. Med Decis Making. 2006;26:565–74. doi: 10.1177/0272989X06295361

28. Kerr KF, Brown MD, Zhu K, et al. Assessing the Clinical Impact of Risk Prediction Models With Decision Curves: Guidance for Correct Interpretation and Appropriate Use. JCO. 2016;34:2534–40. doi: 10.1200/JCO.2015.65.5654

29. R: The R Project for Statistical Computing. https://www.r-project.org (accessed 24 Sept 2026).

30. Sjoberg D. dcurves: Decision Curve Analysis for Model Evaluation. R package version 0.5.0. https://www.danieldsjoberg.com/dcurves/ (accessed 24 Sept 2026).

31. Wickham H, François R, Henry L, et al. dplyr: A Grammar of Data Manipulation. R package version 1.1.4, https://dplyr.tidyverse.org (accessed 24 Sept 2026).

32. Sievert C. Interactive Web-Based Data Visualization with R, plotly, and shiny. https://plotly-r.com (accessed 24 Sept 2026).

33. Belsher BE, Smolenski DJ, Pruitt LD, et al. Prediction Models for Suicide Attempts and Deaths: A Systematic Review and Simulation. JAMA Psychiatry. 2019;76:642. doi: 10.1001/jamapsychiatry.2019.0174

34. Kessler RC, Bossarte RM, Luedtke A, et al. Suicide prediction models: a critical review of recent research with recommendations for the way forward. Mol Psychiatry. 2020;25:168–79. doi: 10.1038/s41380-019-0531-0

35. Seyedsalehi A, Fazel S. Suicide risk assessment tools and prediction models: new evidence, methodological innovations, outdated criticisms. BMJ Ment Health. 2024;27:e300990. doi: 10.1136/bmjment-2024-300990

36. Kline-Simon AH, Sterling S, Young-Wolff K, et al. Estimates of Workload Associated With Suicide Risk Alerts After Implementation of Risk-Prediction Model. JAMA Netw Open. 2020;3:e2021189. doi: 10.1001/jamanetworkopen.2020.21189

37. McCutchan PK, Yates BT, Jobes DA, et al. Costs, benefits, and cost-benefit of Collaborative Assessment and Management of Suicidality versus enhanced treatment as usual. Durand-Zaleski I, editor. PLoS ONE. 2022;17:e0262592. doi: 10.1371/journal.pone.0262592

38. Wynants L, Van Smeden M, McLernon DJ, et al. Three myths about risk thresholds for prediction models. BMC Med. 2019;17:192. doi: 10.1186/s12916-019-1425-3

39. Goldstein Grumet J, Boudreaux ED. Universal Suicide Screening Is Feasible and Necessary to Reduce Suicide. PS. 2023;74:81–3. doi: 10.1176/appi.ps.202100625

40. Obermeyer Z, Powers B, Vogeli C, et al. Dissecting racial bias in an algorithm used to manage the health of populations. Science. 2019;366:447–53. doi: 10.1126/science.aax2342

