## Supplemental File 1 for "Considerations for implementing a machine learning model for suicide risk in the health systems that serve American Indian communities"

### Supplemental File 1: IHS Sample Analyses

#### **FIGURES: CALIBRATION PLOTS IN IHS-VISIT COHORT**

**Figure S1 *–*** *Calibration Plot of Raw and Isotonic Regression Calibrated Vanderbilt Model across all Visits Between October 2019 and October 2021 (N = 11,842 patients; 148,914 visits)*

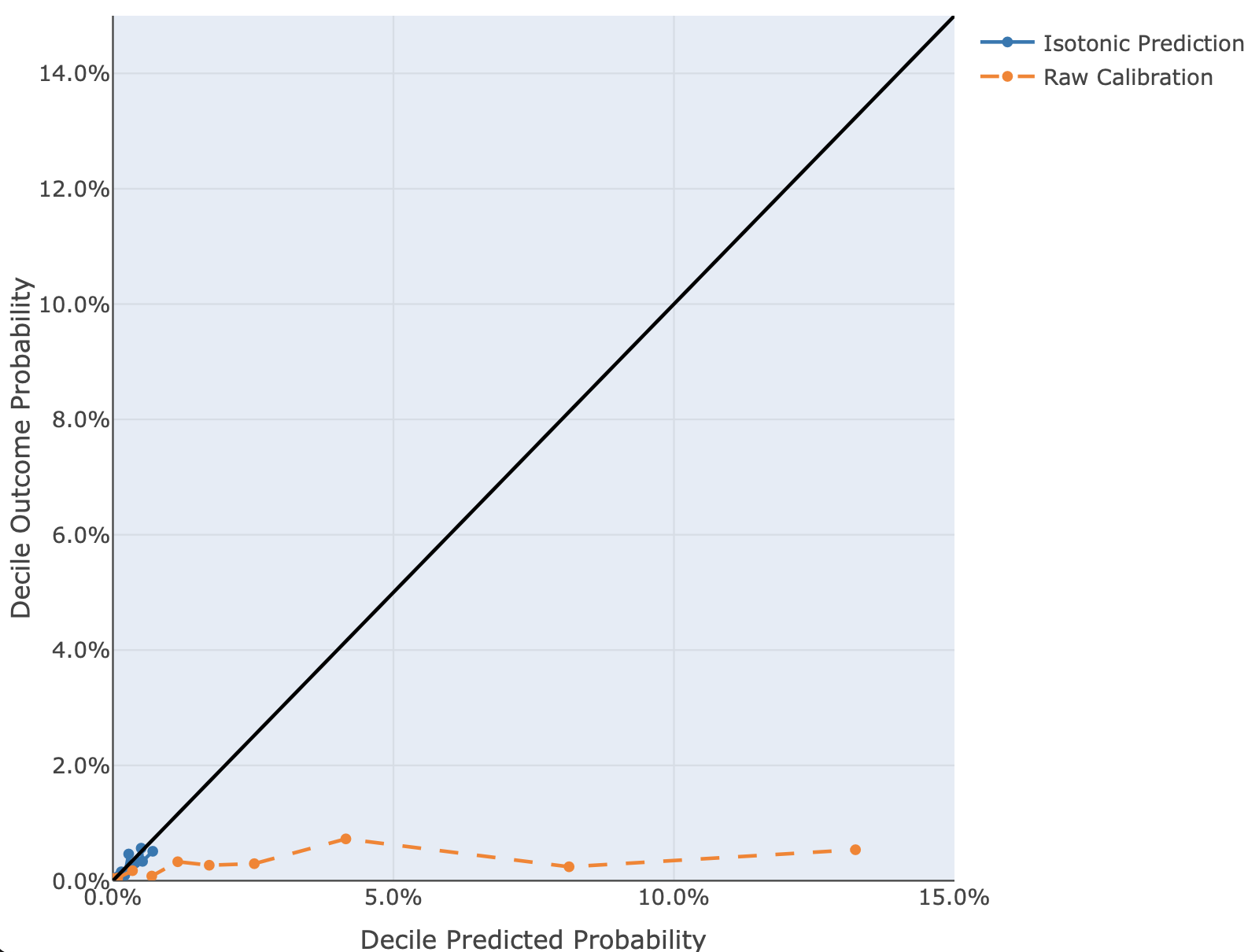

**Figure S2 –** *Calibration Plot of Raw and Isotonic Regression Calibrated Mental Health Research Network (MHRN) Model across all Visits Between October 2019 and October 2021 (N = 11,842 patients; 148,914 visits)*

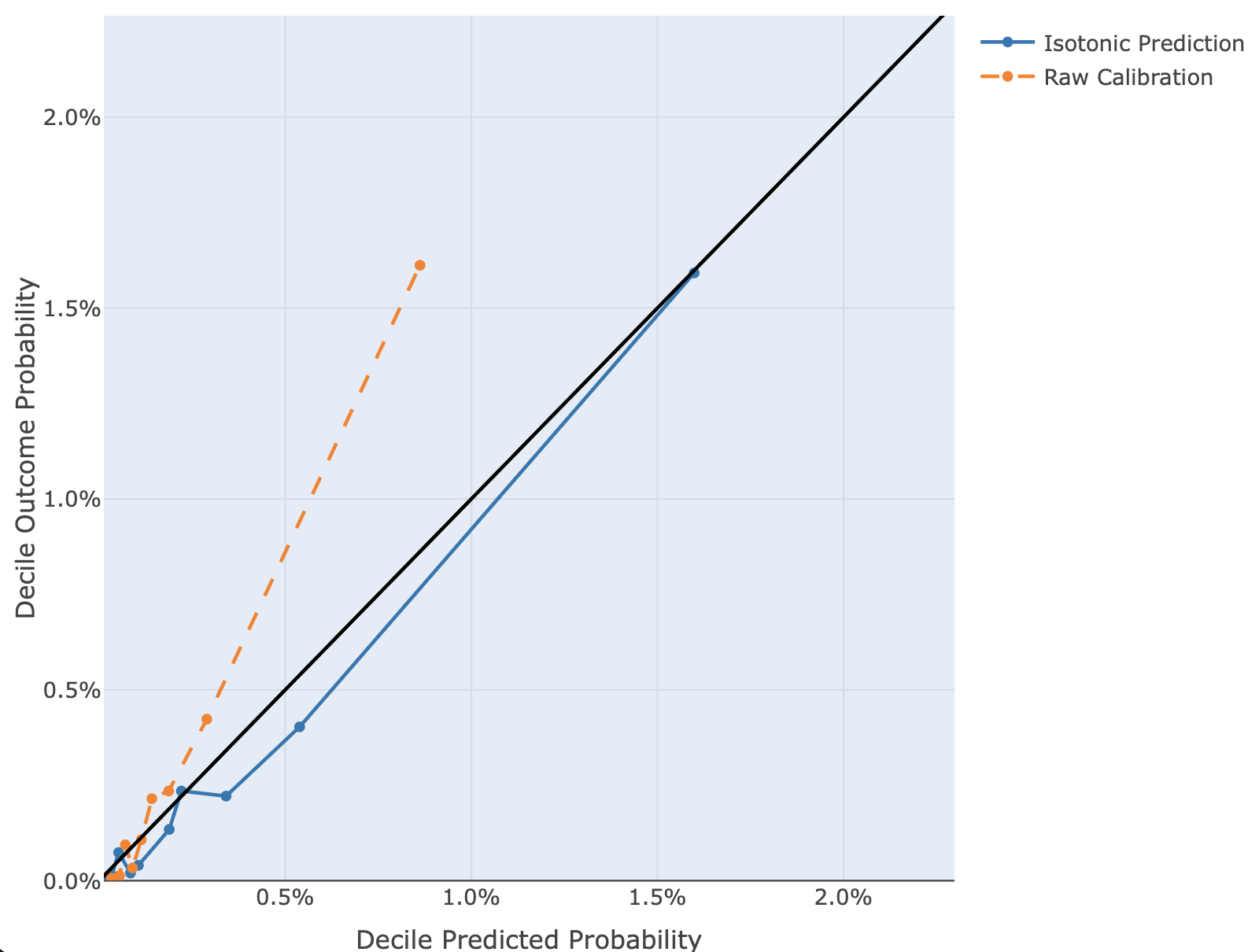

**Figure S3 *–*** *Calibration Plot of Raw and Isotonic Regression Calibrated IHS Random Forest (RF) Model across all Visits Between October 2019 and October 2021 (N = 11,842 patients; 148,914 visits)*

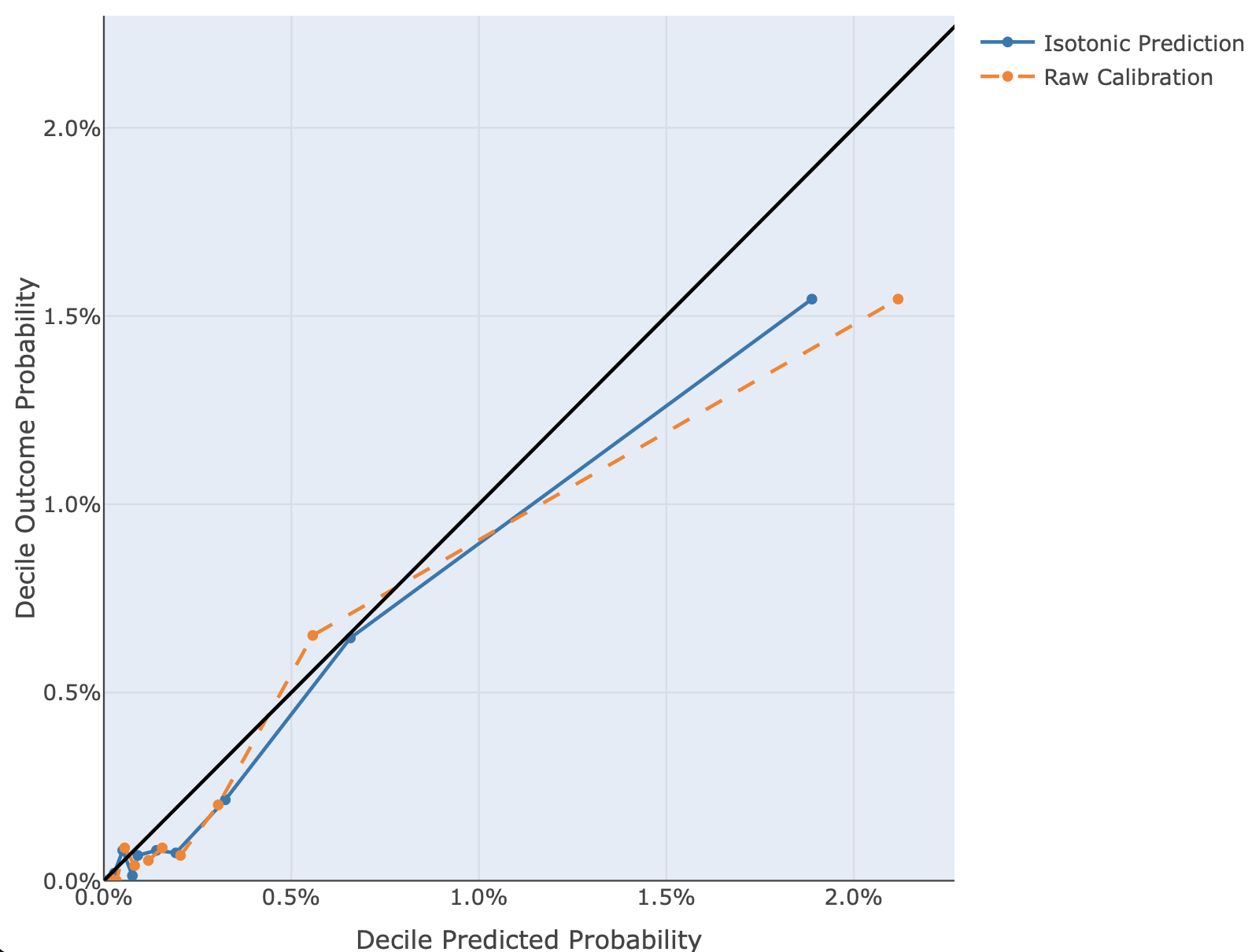

**Figure S4 *–*** *Calibration Plot of Raw and Isotonic Regression Calibrated IHS Logistic Regression (LR) Model across all Visits Between October 2019 and October 2021 (N = 11,842 patients; 148,914 visits)*

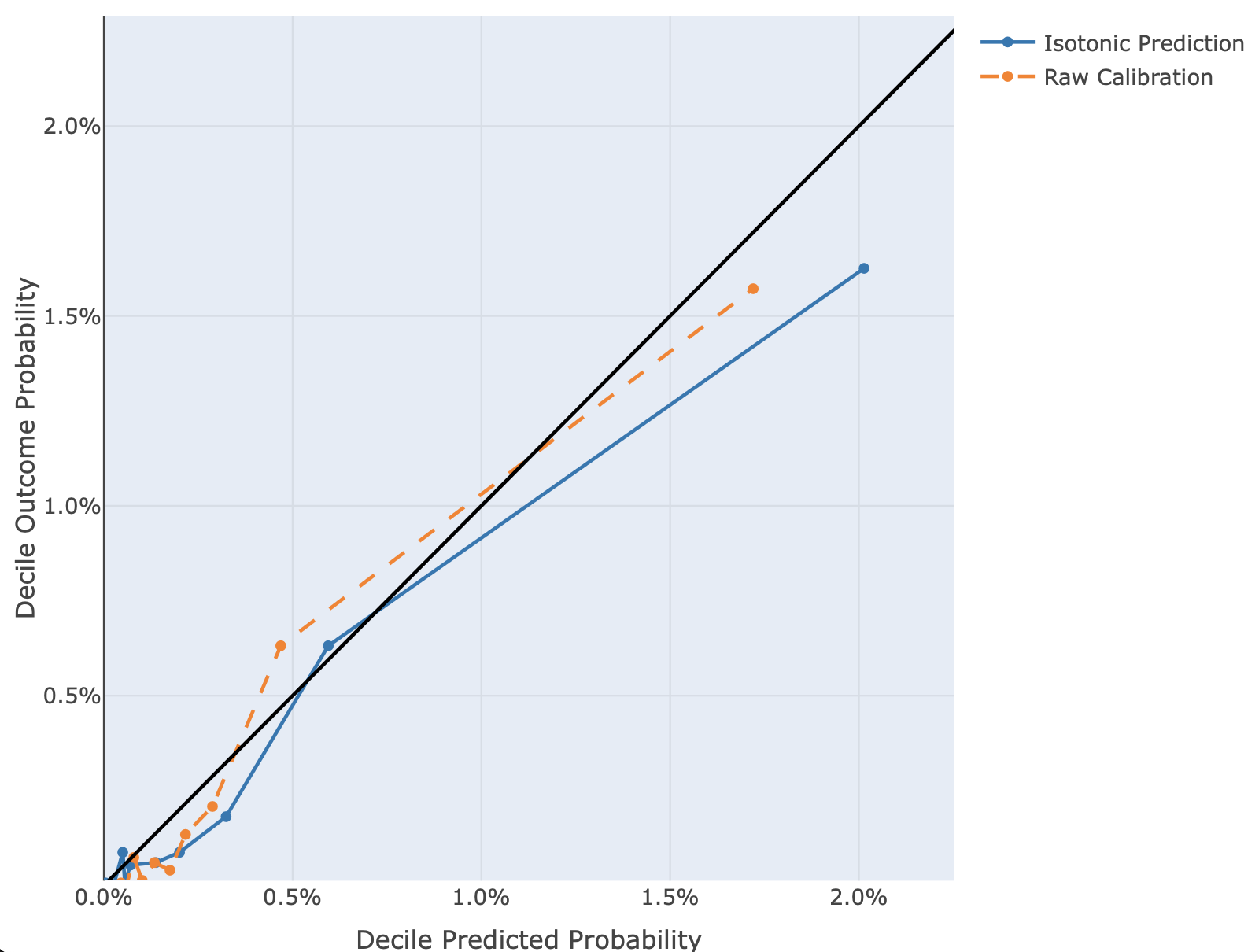

#### **TABLES OF IHS-VISIT COHORT**

**Table S1 *–*** *Sample Demographic Characteristics of Patients With One or More IHS Visits Between October 2019 and October 2021 (N = 11,842 patients; 148,914 visits)*

| **Characteristics** | **N = 11,842** |
| --- | --- |
| *Sex, n (%)* |  |
| Female | 6,190 (52.3) |
| Male | 5,651 (47.7) |
| Unknown | 1 (<0.1) |
| *Age at first visit, median (IQR)* | 38 (28, 55) |
| *Ethnicity, n (%)* |  |
| Hispanic, Latino, or Spanish origin | 88 (0.7) |
| Not Hispanic, Latino, or Spanish Origin | 11,606 (98.0) |
| Unknown | 148 (1.3) |
| *Race, n (%)* |  |
| American Indian or Alaska Native | 11,226 (94.8) |
| Asian | 37 (0.3) |
| Black/African-American | 17 (0.1) |
| Native Hawaiian/Pacific Islander | 11 (<0.1) |
| White | 362 (3.1) |
| Other | 70 (0.6) |
| Unknown | 118 (1.0) |
| *Health Insurance at first visit, n (%)* |  |
| Not enrolled | 2,437 (20.6) |
| Medicare | 1,818 (15.4) |
| Medicaid | 7,574 (64.0) |
| Private Insurance | 1,303 (11.0) |
| *Suicide outcomes, n (%)* |  |
| Attempted suicide | 128 (1.1) |
| Died by suicide | 17 (0.1) |
| Attempted or died by suicide | 144 (1.2) |
| Attempted or died by suicide within 90 days of last visit | 102 (0.9) |
| *Existing screening, n (%)^1^* | 1,012 (8.5) |
| ^1^ Existing screening refers to a positive Ask Suicide-Screening Questions (ASQ) survey result, a positive risk assessment via the Columbia-Suicide Severity Rating Scale (C-SSRS), a diagnosis of ideation within 90 days of an included visit, or any history of a previous suicide attempt. Existing screening outcomes before 10/1/19 were censored. | |

**Table S2 *–*** *Standardized Net Benefit, Summary Statistics, and Predicted Advantages of the IHS Logistic Regression (IHS-LR) Model in Comparison Existing Screening Among IHS Visits Between October 2019 and October 2021 (N = 11,842 patients; 148,914 visits)*

| Percentile |  | Standardized Net Benefit^1^ | | IHS-LR Model | | | Comparing IHS-LR Model to Existing Screening | |
| --- | --- | --- | --- | --- | --- | --- | --- | --- |
|  | p(t)^2^ (%) | Existing Screening^3^ (%) | IHS-LR Model (%) | PPV (%) | Sensitivity (%) | Specificity (%) | Increase in true positives per 1,000 visits with no increase in false positives^4^ | Percent increase in true positive cases with no increase in false positive cases^5^ |
| >50 | 0.09 | 41.94 | 75.89 | 0.48 | 96.21 | 44.20 | 0.93 | 33.94 |
| >75 | 0.34 | 36.56 | 50.00 | 0.86 | 85.33 | 72.92 | 0.37 | 13.44 |
| >90 | 0.82 | 26.16 | 26.20 | 1.32 | 65.60 | 86.66 | <0.01 | 0.04 |
| >95 | 1.21 | 17.63 | 20.96 | 2.04 | 46.34 | 94.02 | 0.09 | 3.34 |
| >98 | 3.12 | -25.14 | 8.09 | 3.72 | 32.92 | 97.77 | 0.91 | 33.23 |
| >99 | 4.17 | -49.37 | -7.14 | 4.31 | 28.20 | 98.25 | 1.16 | 42.23 |
| ^1^ Standardized Net Benefit presented as the average SNB across 10 folds  ^2^ p(t): Threshold Probability  ^3^ Existing screening: Suicide screening approach including the Ask Suicide-Screening Questions (ASQ) survey, risk assessment via the Columbia-Suicide Severity Rating Scale (C-SSRS), diagnosis of ideation within 90 days of an included visit, or any history of a previous suicide attempt. Existing screening had a PPV of 2.01%, a sensitivity of 43.87%, and a specificity of 94.11%.  ^4^ The hypothesized increase in true positive cases per 1,000 visits was calculated as the net benefit at the specified p(t) of the IHS-LR model subtracted by the net benefit at the specified p(t) of existing screening.  ^5^ The percentage increase in true positive visits flagged with no percent increase in false positive visits flagged was calculated as the standardized net benefit at the specified p(t) of the IHS-LR model subtracted by the standardized net benefit at the specified p(t) of existing screening. | | | | | | | | |

#### **FIGURES: IHS-VISIT COHORT**

**Figure S5** *– Comparison of Standardized Net Benefits for the Identification of Patients Who Had a Suicide Attempt or Died by Suicide Within 90 Days of IHS Visits Between October 2019 and October 2021 (N = 11,842 patients; 148,914 visits)*

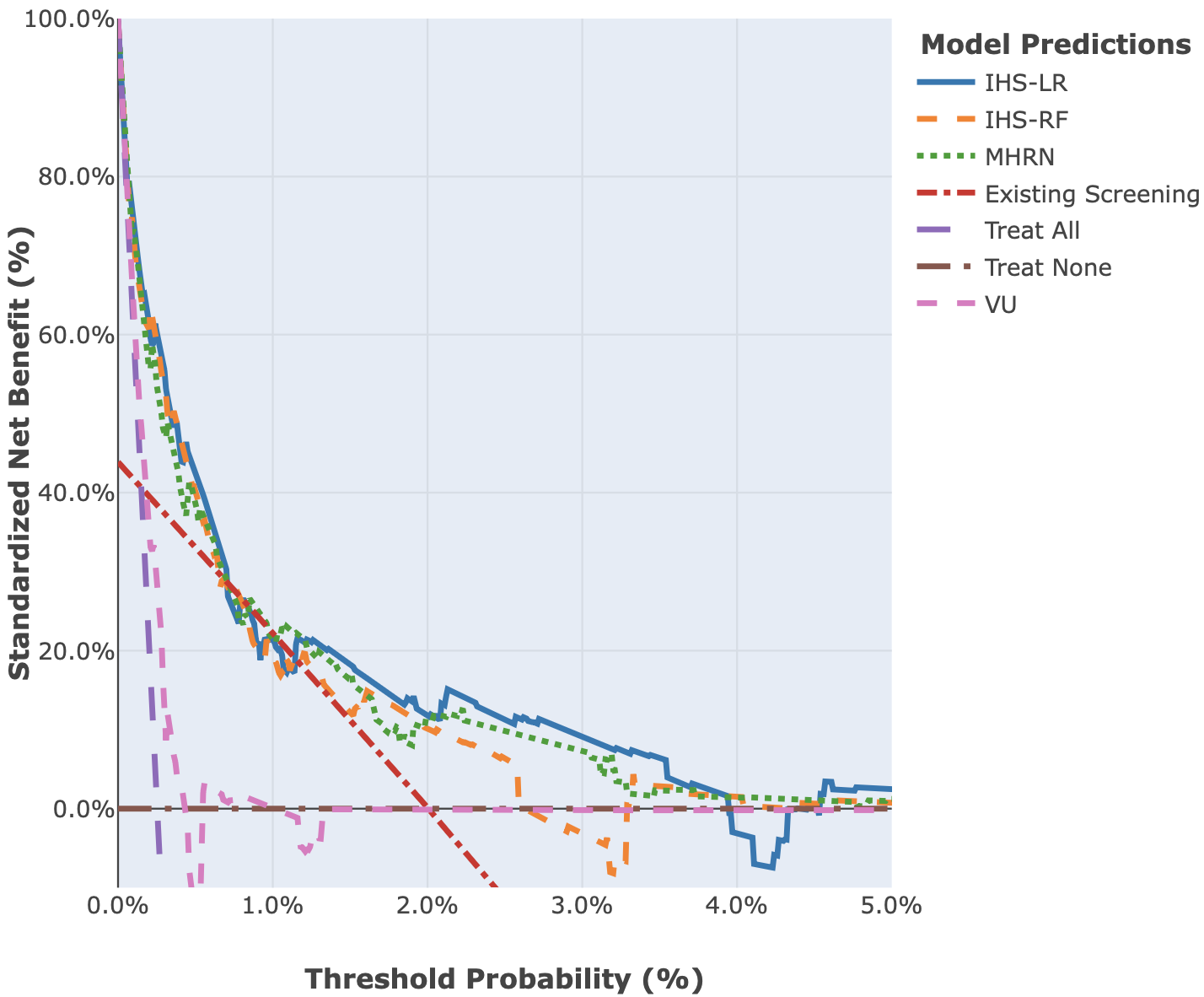

**Figure S6 *–*** *Workload Estimates in Percentage of Alerts on IHS Visits per Week Comparing the Native-RISE IHS-LR Model to Existing Screening Between October 2019 and October 2021 (N = 11,842 patients; 148,914 visits)*

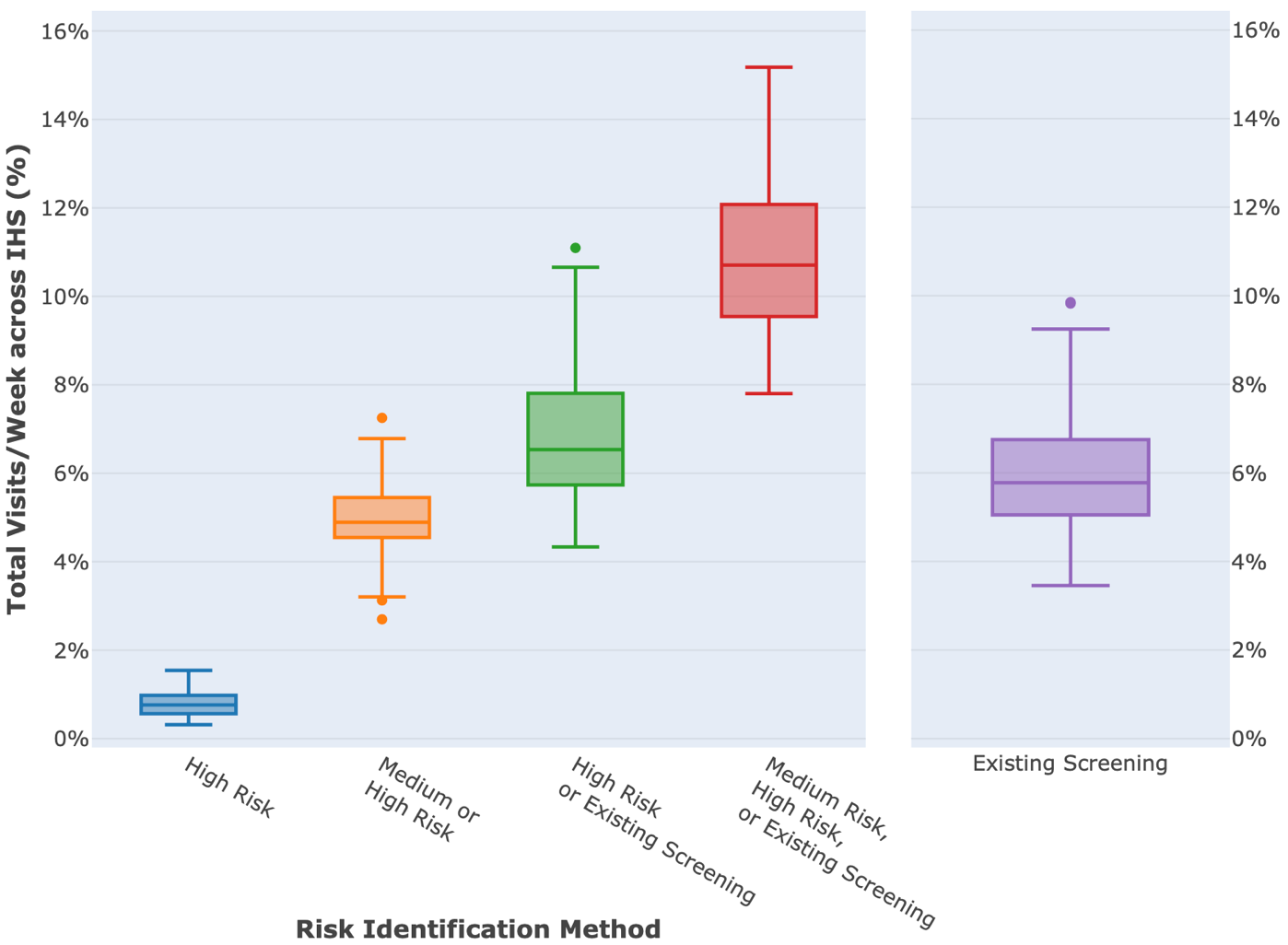
