## Supplemental File 2 for "Considerations for implementing a machine learning model for suicide risk in the health systems that serve American Indian communities"

### Supplemental File 2: Calibration Plots of Ed-Visit Cohort

**Figure S1 *–*** *Calibration Plot of Raw and Isotonic Regression Calibrated Vanderbilt Model across all ED Visits Between October 2019 and October 2021 (N = 9,244 patients; 42,915 visits)*

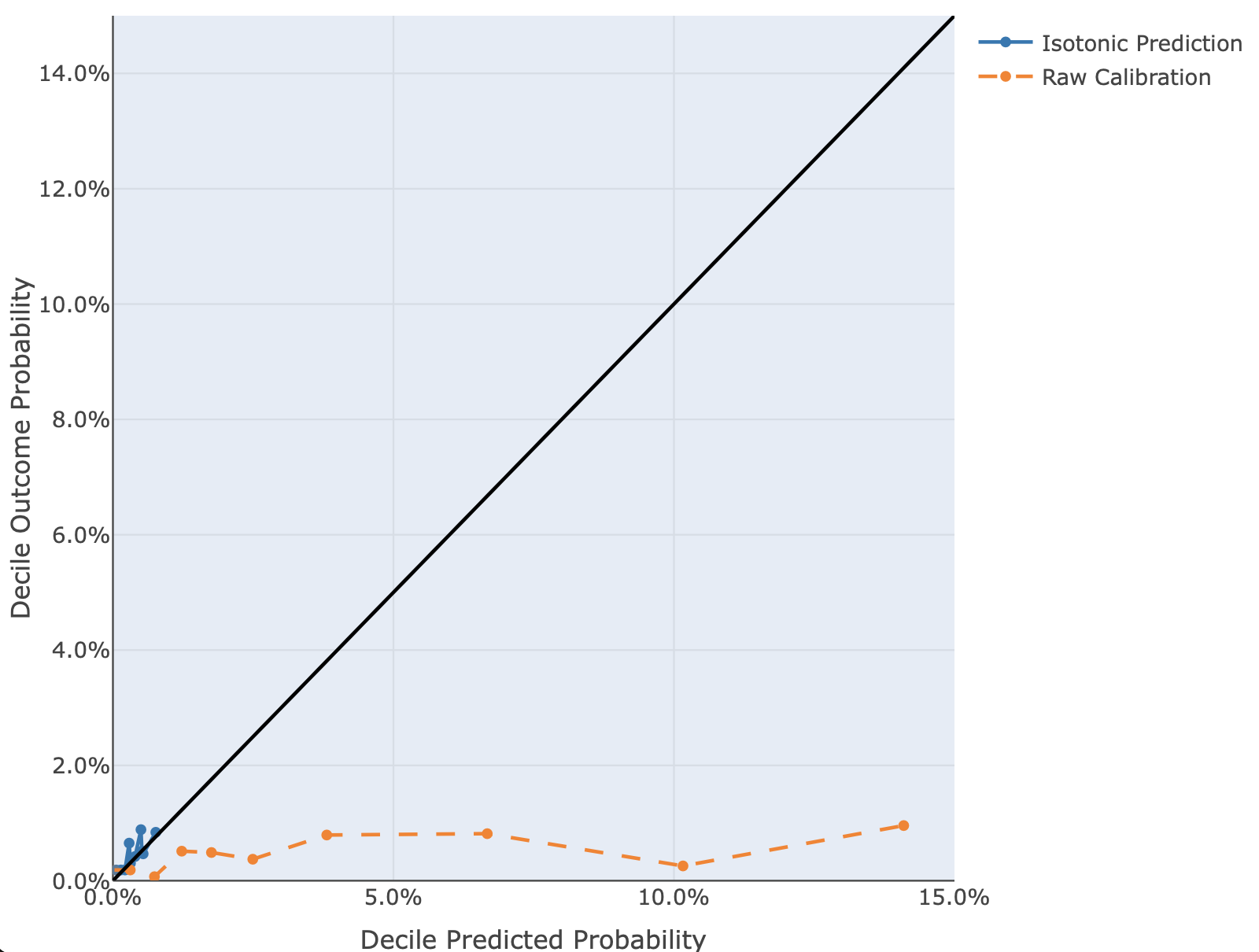

**Figure S2 *–*** *Calibration Plot of Raw and Isotonic Regression Calibrated Mental Health Research Network (MHRN) Model across all ED Visits Between October 2019 and October 2021 (N = 9,244 patients; 42,915 visits)*

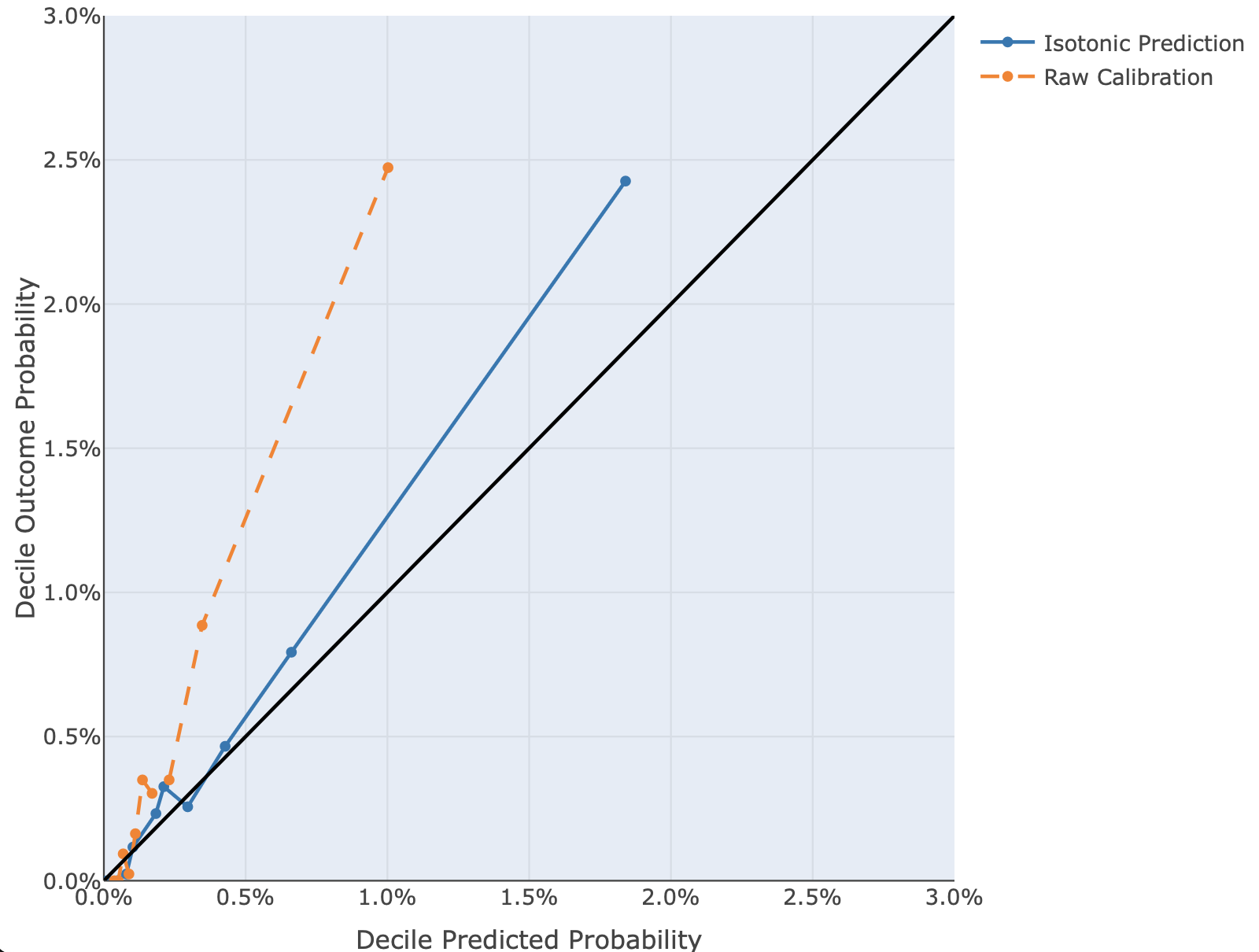

**Figure S3 *–*** *Calibration Plot of Raw and Isotonic Regression Calibrated IHS Random Forest (RF) Model across all ED Visits Between October 2019 and October 2021 (N = 9,244 patients; 42,915 visits)*

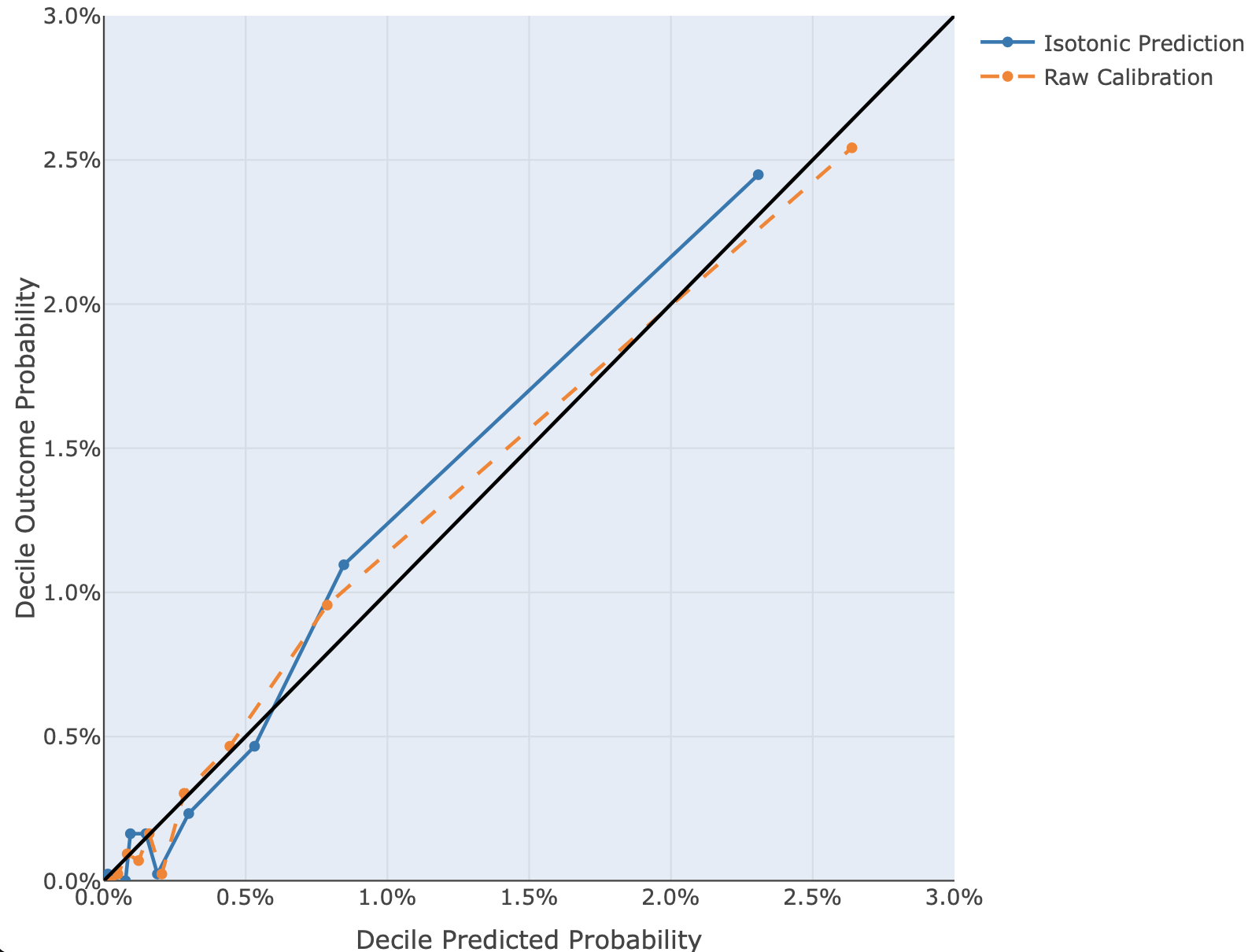

**Figure S4 *–*** *Calibration Plot of Raw and Isotonic Regression Calibrated IHS Logistic Regression (LR) Model across all ED Visits Between October 2019 and October 2021 (N = 9,244 patients; 42,915 visits)*

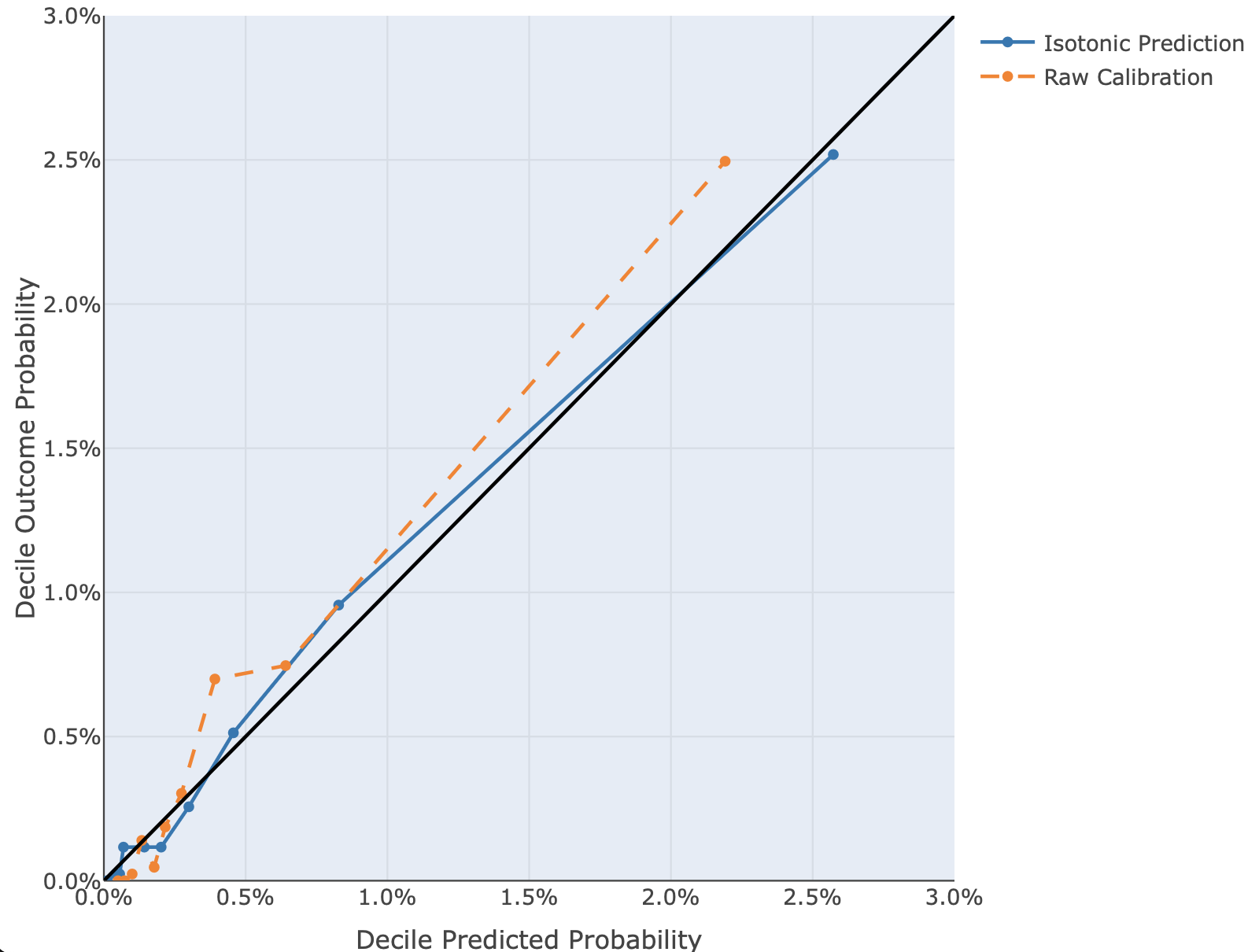
