## Supplemental File 3 for "Considerations for implementing a machine learning model for suicide risk in the health systems that serve American Indian communities"

### Supplemental File 3: Additional Methods

#### **Net benefit: Threshold Probability Explanation**

Threshold probability is defined as the point at which the expected benefit of treatment is equivalent to the potential harm of unnecessary treatment. When operationalizing a predictive model at the threshold probability, any predicted probability greater than the threshold probability would be classified as positive (signifying that the expected benefits outweigh potential harms), while predicted probabilities below the threshold would be classified as negative (signifying that the potential harms outweigh expected benefits).^1^ However, threshold probability is difficult to quantify on a population level due to patient-specific harms or benefits, such as the efficacy of treatments available, the resources needed to provide treatment, or the varying harms associated with misclassification.[1]

#### **Net benefit: Example Interpretation of Probability Threshold and Harm-to-Benefit Ratios**

If a provider always treated a patient with a 2% risk of a suicide attempt or death in the next 90 days, the net benefits can be compared across all models by examining the number of true positives and false positives when 2% is used as the probability threshold. Further, probability thresholds can be interpreted as "harm-to-benefit" ratios – the number of patients that providers are willing to assess, to identify one patient truly at risk of suicide.[2] The odds of the probability threshold is equivalent to the risk/benefit tradeoff. For example, if a clinician preferred to assess no more than 20 patients to identify one patient truly at-risk, they equivalently would prefer a risk/benefit tradeoff of 1:19 or a probability threshold of 5%.

#### **Net benefit: Discussion of Interpretability in the Context of Suicide-Related Care**

Engaging providers to select meaningful threshold probabilities proved challenging. Providers consulted to interpret these decision curves generally conceptualized risk on a 0-100% scale, while smaller increments of risk relevant to suicide risk models (0-5%) were more challenging to discern between. This limited our ability to establish meaningful harm-to-benefit ratios that corresponded with the sensitivity. For example, the high-risk threshold was set to have a sensitivity that approximated the sensitivity of existing screening, and corresponded with a probability threshold of approximately 1.6%, a workload estimate of approximately 1.3% visits per week identified, and a sensitivity of 42.5%. However, the harm-to-benefit ratio associated with the high-risk threshold of 1:62 was not as interpretable to clinicians as other estimates provided, such as the workload estimate.

### References

1. Vickers AJ, Elkin EB. Decision Curve Analysis: A Novel Method for Evaluating Prediction Models. Med Decis Making. 2006;26:565–74. doi: https://doi.org/10.1177/0272989X06295361
2. Van Calster B, Wynants L, Verbeek JFM, et al. Reporting and Interpreting Decision Curve Analysis: A Guide for Investigators. European Urology. 2018;74:796–804. doi: https://doi.org/10.1016/j.eururo.2018.08.038
